# DETERMINANTS OF HEALTH SEEKING BEHAVIOUR OF NOMADIC PASTORALISTS IN LOW AND MIDDLE-INCOME COUNTRIES: A SCOPING REVIEW

**DOI:** 10.1101/2025.07.16.25331642

**Authors:** James Ngeruro Kariuki

**Affiliations:** A2108D12651553, Dissertation Project (48221), LJMU-7506-PUBUNI-48221

**Keywords:** health-seeking behaviours, pastoralists, nomadic, healthcare services, traditional methods, sociocultural factors

## Abstract

Access and utilization of healthcare facilities have been an important factor in determining the health and wellbeing gap in society. However, these opportunities have not been benefiting some communities, such as the pastoralist communities, due to the constant movement in search of pasture for the animals. These communities are also faced with other issues, such as language and cultural concerns, which may impact how they interact with healthcare providers. In that regard, this review was carried out to assess some of the issues that impact the health-seeking behaviours of these communities in order to understand the underutilization of healthcare facilities.

In meeting the research, the study adopted a scoping review in which the researcher relied on 25 sources to provide evidence on the issues. These articles were sourced from Google Scholar, Scopus PubMed, and ScienceDirect.

The study indicated that health-seeking behaviours have been impacted by negative perceptions, misinformation, misconceptions regarding the mainstream care system and increased institutional barriers. Cultural attitudes and low knowledge and education level helps in sustaining the negative perception that later shaped the health-seeking behaviours.

Misinformation also created misconception around various diseases and symptoms, thereby limiting the urgency of seeking care from qualified medical practitioners. Distance was a significant factor because it made the accessibility of healthcare facility impossible. The study further recommends the improvement of knowledge through awareness to minimize misconceptions about health or diseases.

## Chapter One: Introduction

### 1.1 Background information

The existence of global discrepancies in healthcare delivery has consistently been attributed to the inequities between affluent and impoverished nations. Lechthaler et al. (2018) argue that the goal of universal healthcare has not been realised due to significant economic challenges, as well as obstacles related to geography, culture, and society. These factors have hindered the government’s efforts to improve health outcomes. Furthermore, according to Wako and Kassa (2017), the limited availability of transportation infrastructure, inadequate service quality, traditional cultural practices, and women’s lack of decision-making power are factors that hinder access to healthcare services and contribute to health disparities. Similarly, Muriithi et al. (2022) suggest that this problem continues to exist in many African countries due to factors such as delayed decision-making in seeking assistance and recognising danger, inadequate service delivery in terms of triage, monitoring, referral, and transportation to and between healthcare facilities.

The matter of health has not only gained attention at the national level but has also become a global problem, leading to its acknowledgement in the global health agenda. Wulifan et al. (2016) highlight that the 2030 sustainable development goals (SDGs) emphasise the importance of reducing maternal mortality and addressing the unmet demand for family planning. These objectives are crucial for promoting the health and well-being of the global population. This would also be crucial in fostering the welfare of communities worldwide. Furthermore, it is a fundamental duty of governments worldwide to ensure the provision of top-notch healthcare services that are accessible, cheap, and inclusive for all residents (WHO, 2018). The World Health Organisation (WHO) has prioritised universal health coverage as its primary objective, encapsulated by the tagline “Health for all - everyone, everywhere” for World Health Day. The attainment of this goal necessitates governments’ commitment to investing in high-quality and easily accessible primary healthcare. Nevertheless, Ali et al. (2019) emphasise the need to direct attention towards marginalised communities, whose progress falls behind that of the overall population, in order to achieve these significant objectives. Duale et al. (2023) contend that despite advancements in achieving universal health care in several Sub-Saharan African nations, the nomadic pastoralist population has often been excluded from these positive developments.

Pastoralists are nomadic individuals whose sustenance primarily relies on cattle, which they periodically or seasonally relocate in pursuit of suitable grazing land and water sources (Duale et al., 2023). The primary demographic consists of nomadic pastoralists who engage in cattle rearing and wander in search of water supplies and grazing lands. They lack permanent settlements and do not own any other resources; instead, they adopt a periodic migration pattern. Africa is home to the largest proportion (60%) of nomadic communities, who have various difficulties in obtaining healthcare services as compared to sedentary groups (Maro et al., 2012). Nomadic pastoralists often reside in border regions, which are characterised by a very unstable environment and are generally inaccessible to official healthcare facilities (Duale et al., 2023). Ali et al. (2019) observe that nomadic tribes in Africa pose challenges to health interventions due to their distant locations, itinerant lifestyles, and linguistic and cultural disparities with established communities. For instance, research conducted in Northern Senegal has shown that the utilisation of malaria prevention measures among nomadic pastoralists is much lower compared to the overall population (Seck et al., 2017). Moreover, in Somalia, nearly 90 per cent of nomads have been documented as being outside the accessibility of the country’s official health facilities (Jillo et al. 2015). According to Gammino et al. (2020), nomadic tribes in Cameroon have been recognised as a possible source of continued transmission of onchocerciasis, a neglected tropical disease that is widespread in many regions of the nation. Consequently, pastoralists have limited access to healthcare services due to various constraints arising from their harsh environment. Additionally, there is a prevailing lack of attention and unequal distribution of healthcare resources among these populations in the country (Montavon et al., 2013). Anthonj et al. (2019) emphasise the importance of the proximity to healthcare facilities, highlighting that these facilities may be far away and challenging to reach, which might impact the behaviour of individuals seeking medical assistance when they are unwell. The treatment or cure of illnesses for people depends on their health-seeking activity. Research on health-seeking behaviour primarily investigates the initial identification of symptoms and tracks the individual’s progression through several phases of understanding and managing their illness. This includes determining whether or not to seek medical attention and actively pursuing health throughout the healing process (Amegbor, 2014). Da Silva et al. (2011) found a clear correlation between healthcare use and the healthcare system of a nation, as well as the available health services. Healthcare use and its patterns are directly influenced by the healthcare-seeking behaviours of individuals. Consequently, it is essential to design and deliver healthcare services by considering data on healthcare-seeking behaviours and utilisation, as well as the various factors that influence them, such as social, economic, physical, cultural beliefs, religious practices, gender norms, and political aspects (Abuduxike et al., 2020). According to Ngwakongnwi (2017), comprehending healthcare-seeking behaviours and their determinants enables government, stakeholders, policy-makers, and health service providers to distribute and oversee available resources effectively, especially in developing nations.

Existing studies have provided important insight into the topic, providing an overview of the possible barriers to positive health-seeking behaviours among pastoralists. These include geographical barriers, cultural and social factors, economic constraints, healthcare infrastructure and education level. Nomadic pastoralists often inhabit isolated and inaccessible regions situated at a considerable distance from healthcare amenities (Wild et al., 2019). The geographical remoteness is a notable obstacle, as it impedes their capacity to get prompt and suitable healthcare treatments. This problem is worsened by the lack of adequate infrastructure and transit choices. Cultural ideas and traditions are essential in influencing the way individuals seek healthcare. The choices of nomadic pastoralists towards obtaining modern healthcare may be influenced by traditional treatment procedures and cultural preferences (Gammino et al., 2020). Moreover, healthcare-seeking behaviour might be influenced by societal norms prevalent in certain communities, whereby factors like gender roles can hinder access to treatments. The economic constraints encountered by nomadic pastoralists are influenced by variables such as poverty and the exorbitant expenses associated with healthcare services (Wulifan et al., 2022). Individuals may be discouraged from obtaining timely and adequate healthcare due to their low financial resources, particularly when they have other financial obligations that compete for their limited income. The lack of sufficient healthcare facilities in the regions inhabited by nomadic pastoralists is a substantial obstacle (Ringo et al., 2018). The absence of healthcare infrastructure, skilled workers, and necessary medical resources might hinder their access to adequate treatment. The correlation between the educational attainment of nomadic pastoralist populations and their propensity to seek healthcare has been noted (Cattaneo, 2019). Increased levels of education are correlated with enhanced health consciousness and a higher probability of using healthcare services.

Gaining insight into these factors is essential for formulating precise treatments and policies designed to enhance the health-seeking conduct of nomadic pastoralists. To tackle these concerns, a comprehensive strategy is needed that takes into account the distinct difficulties presented by their itinerant way of life and the particular cultural environments in which they reside. These findings may be used by researchers and policymakers to develop efficient and culturally attuned healthcare policies specifically designed for nomadic pastoralist groups in low and middle-income nations.

### 1.2 Theoretical Framework

The theory that is important and relevant to this review and the discussion of its findings is Anderson’s behavioural model. In essence, Andersen’s behavioural model for health services utilisation offers a conceptual framework for comprehending the accessibility and utilisation of health services, as well as identifying the determinants that influence an individual’s choice to use or abstain from using available health services (Andersen, 1995). According to this behavioural model, a person’s utilisation of health services is influenced by a series of variables, including predisposing, enabling, and need factors. Andersen focuses his efforts on constructing a model that identifies factors that either facilitate or hinder the use of healthcare services by individuals. This method is often used to uncover variables that are linked to the use of health care. The model described is a comprehensive, multilevel model that incorporates both personal and situational aspects that contribute to the utilisation of health care. This model has gained widespread acceptance and use in industrialised countries (Andersen et al., 2011). The model provides a comprehensive analysis of health care use by patients, effectively integrating many elements and offering scientists a holistic explanation of this phenomenon (Holde et al., 2018). The primary objective of the Model is to comprehend the motivations and factors behind individuals’ utilisation of healthcare services, evaluate disparities in the availability of health services, and contribute to the formulation of policies that promote fair and equal access to care (Travers et al., 2020). The model is relevant in the context of this study as it would help describe the predisposing factors, enabling factors, and need factors that have been influencing the health-seeking behaviours of the Nomad Pastoralists and the subsequent use of their healthcare services.

### 1.3 Statement of the Problem

According to Lechthaler et al. (2018), in many low- and middle-income countries (LMICs) with nomadic pastoralist populations, access to quality healthcare is often determined by factors such as ethnicity and religious affiliation rather than solely by poverty levels. These populations are typically geographically isolated and have political systems based on clientelism and sectarian structures. Healthcare inequality, also known as healthcare disparities, pertains to the variation in health and healthcare standards across different racial and ethnic groups, sexual orientations, and financial gaps (Ali et al., 2019). The health results of these communities, which consist of nomadic pastoralists, are inferior and marginalised in comparison to the mainstream population due to their vulnerability. Over 70% of women in predominately pastoralist areas of Ethiopia rely on traditional birth attendants (TBAs) for support during childbirth at home. This practice contributes to a higher incidence of maternal mortality among the pastoralist population (Ayalew et al., 2022). Furthermore, among the African populations, nomadic pastoralists in Africa exhibit elevated rates of morbidity and death for many avoidable ailments (Kenya National Bureau of Statistics, 2015). This highlights the need to enhance health outcomes by evaluating the reasons that hinder these women from seeking assistance from experienced healthcare professionals. Anthonj et al. (2019) suggest that studying the behaviour and decision-making of communities affected by ill-health might provide useful insights for designing future health treatments and managing wetlands in a way that promotes health.

### 1.4 Study Aims

The study aims to identify nomadic pastoralists’ health-seeking behaviours by assessing the literature on the issues they face while trying to access quality care services in LMICs.

### 1.5 Research Objectives

1. To assess the issues limiting the health-seeking behaviours of the nomadic pastoralists’ in LMICs
2. To assess the impact of low health-seeking behaviours in the acquisition of health service delivery and health the overall health outcomes of the nomadic pastoralists in LMICs.
3. To provide recommendations on the best practices that can be adopted to promote health-seeking behaviours among the nomadic pastoralists in LMICs.

### 1.6 Contribution

The study results might guide the creation of focused healthcare policies and treatments that are specially customised to meet the requirements of nomadic pastoralist communities. Policymakers may use these insights to develop solutions that effectively tackle the distinct obstacles presented by individuals’ mobile lives, geographical seclusion, and cultural convictions. The study aims to identify the primary obstacles that hinder nomadic pastoralists’ access to healthcare. This information will be used to develop strategies to enhance the accessibility and availability of healthcare services for this population. This might include the creation of mobile healthcare facilities, enhanced infrastructure for transportation, and the education of healthcare workers to efficiently operate in distant regions. Gaining insight into the cultural and socioeconomic determinants that impact individuals’ decisions to seek healthcare is essential for developing healthcare treatments that are culturally attuned. The study has the potential to enhance the development of methodologies that acknowledge and integrate ancient healing traditions while also addressing gender-specific issues, therefore enhancing the acceptance of contemporary healthcare services.

### 1.7 Dissertation Structure

This dissertation is structured into five major chapters. The first chapter has provided the background information on the topic, including research objectives and theoretical framework. The second chapter is the methodology section, justifying the approaches used in data collection and analysis. This chapter also provides a detailed account of the search strategy adopted in getting the rights articles for the final analysis. The third chapter provides findings from the selected sources in relation to the issues being investigated. The fourth chapter is the discussion section on major themes and how the findings met the study objectives. This chapter also provides the implication of these findings to practices and public health theory. The last chapter is the conclusion of the findings, including recommendations for policy and future research on the topic.

## Chapter Two: Methodology

### 2.1 Research question

As indicated earlier, the main aim of the study was to assess the health-seeking behaviours of Nomadic pastoralists in LMICs. In that regard, the research question guiding this review is, “What factors influence the health-seeking behaviours of nomadic pastoralists in low and middle-income countries?” such assessment would be considered important in developing interventions that can enhance the access of this population to healthcare services.

### 2.2 Study Design

The current research adopts a systematic review approach as it seeks to map evidence through the literature on the factors affecting the health-seeking behaviours among the nomadic pastoralists in the LMICs. A scoping review is a thorough technique for synthesising current literature on a specific issue, providing a beneficial study design that is essential for research and evidence-based practice (Munn et al. 2018). This becomes important in health research as it helps provide evidence that can be used to promote health among the population being investigated (nomadic pastoralists). The significance of scoping review further arises from its capacity to map the terrain of available information, offering a comprehensive picture of research on a given topic. This is not only important in outlining verified interventions but also in providing gaps regarding the health issue for further research. Scoping reviews allow researchers to thoroughly examine the sources and forms of evidence relevant to a certain study field in order to assess the present state of knowledge about the issue (Wulifan et al. 2016). Unlike systematic reviews, which concentrate on a specific research issue, scoping reviews seek to uncover essential ideas, evidence gaps, and methodological variances within a larger subject area.

The fundamental argument or rationale for adopting a scoping review is its capacity to handle complicated and varied research issues, particularly when the current literature is extensive and heterogeneous (Hanelt et al., 2021). Researchers may assess the breadth, range, and character of available information by methodically mapping the current literature, making it easier to identify gaps and areas that need future inquiry (Peters et al., 2021). This is especially useful in new or transdisciplinary domains, where a more comprehensive comprehension of the current knowledge base is required. Scoping reviews also help to build research agendas by exposing regions with inadequate data and encouraging academics to concentrate on certain characteristics or gaps in a subject (Lau et al., 2015). They also help policymakers and practitioners make informed choices by synthesising and presenting available information in an understandable format.

Furthermore, the inclusivity aspect of scoping reviews enables the integration of a variety of research designs, procedures, and sources, encouraging a comprehensive understanding of the issue under examination (Pham et al., 2014). This inclusion is especially useful when dealing with complicated topics that a single research design cannot properly address.

### 2.3 The search Strategy

The search strategy involved the identification of databases and the development of key terms to improve the identification of relevant studies on health-seeking behaviours among nomadic pastoralist communities in the low and middle income countries. The databases considered in this case include PubMed, Google Scholar, Scopus and ScienceDirect. The search terms and phrases used include “nomadic pastoralists”, “nomadic communities”, “health-seeking behaviour”, “determinants healthcare”, “institutional barriers to healthcare”, “low and middle-income countries”, “developing countries”, “mobile health services”, “cultural beliefs and healthcare,” “access to healthcare services”, “health disparities” “sociocultural factors healthcare services” and “economic influences on health-seeking behaviour”. The review also included Boolean operators (AND, OR) to improve the outcomes of the search. This led to the following search terms: “Nomadic pastoralists AND health-seeking behaviour”, “Determinants of health-seeking behaviours AND nomadic communities OR nomadic populations OR nomadic pastoralists AND low and middle-income countries OR developing countries”, “Economic influences OR determinants AND health-seeking behaviour AND nomadic pastoralists”, “Environmental determinants OR Health education OR Cultural beliefs OR Institutional barriers OR Sociocultural factors AND healthcare services AND nomadic communities.”

### 2.4 Inclusion and Exclusion Criteria

In developing the eligibility criteria, the PICOS (Population Intervention/Issue Comparison, Outcomes Study design) framework was vital for this research, leading to the inclusion and exclusion criteria. In terms of the population, studies focus on nomadic pastoralists as the primary population, while others focus on other people rather than nomadic pastoralists. The relevance of this is to get studies whose focus is on the targeted population relevant to the development of the study. In terms of the issue or intervention, studies focusing on health-seeking behaviours will be included, whereas those focusing on other issues, such as treatment mode, are excluded from the review. The relevance of focusing on studies on determinants of health-seeking behaviours is to have those studies that are relevant and aligned to the research question. In terms of comparison, the studies selected are those that provided a comparison of how different determinants impact health-seeking behaviours. The rationale of this is to increase the in-depth assessment of different factors influencing these behaviours among the pastoralist communities.

Another concern is the outcomes of these studies in which the researcher focuses on those that focus on the determinants of the factors influencing health-seeking behaviour. These include sociocultural factors, economic factors, distance to the health facilities, and institutional barriers, among others. This is important in aligning the outcomes to the specific issues being investigated in this review. The review includes mixed methods studies, qualitative research and quantitative research. This is important in allowing the review to be based on a wide range of methods to provide evidence based on the primary research. The research also includes peer-reviewed and published studies while excluding company reports, dissertations and news articles. The relevance of this is to provide reliable information or data on the health-seeking behaviours among the nomadic pastoralist communities.

**Table 1:** Inclusion and Exclusion Criteria.

| Criteria | Inclusion | Excluded |
| --- | --- | --- |
| Study designs | Mixed methods research, qualitative design and quantitative research designs | Systematic reviews and scoping reviews |
| Location of the study selected | Low and middle-income countries in Asia, the Caribbean and Sub-Saharan Africa (SSA) | High-income countries |
| Date of the research and its publication | 2013 to 2024 | Before 2013 |
| Language | Studies published in the English language | Studies published in any other languages. |
| Research | Research focusing on determinants of health-seeking behaviours among pastoralist communities | Studies focus on other communities rather than pastoralists. |

### 2.5 Study selection

The screening method followed PRISMA criteria, which are an evidence-based approach for reporting systematic reviews (including designs such as meta-analyses) (PRISMA, 2021; Page et al., 2021), as seen in the flowchart below. The first step was to identify general results after using the keyword alone, which produced more hits, but the researcher narrowed the search further by incorporating a time limit for publishing and the Boolean operators AND/OR, which reduced the number to a manageable figure of 414. Before screening, 206 duplicate records were deleted from the 414 recognised articles, leaving just 208 items to be examined. After adopting the inclusion and exclusion parameters, particularly the usage of titles and abstracts that were irrelevant to the review, 161 records were eliminated, leaving 47 reports for retrieval. However, 4 records were not obtained, resulting in 43 reports being evaluated for eligibility. After removing 18 publications that focused on the general population rather than pastoralists, a total of 25 articles were considered for evaluation.

**Figure 1:**
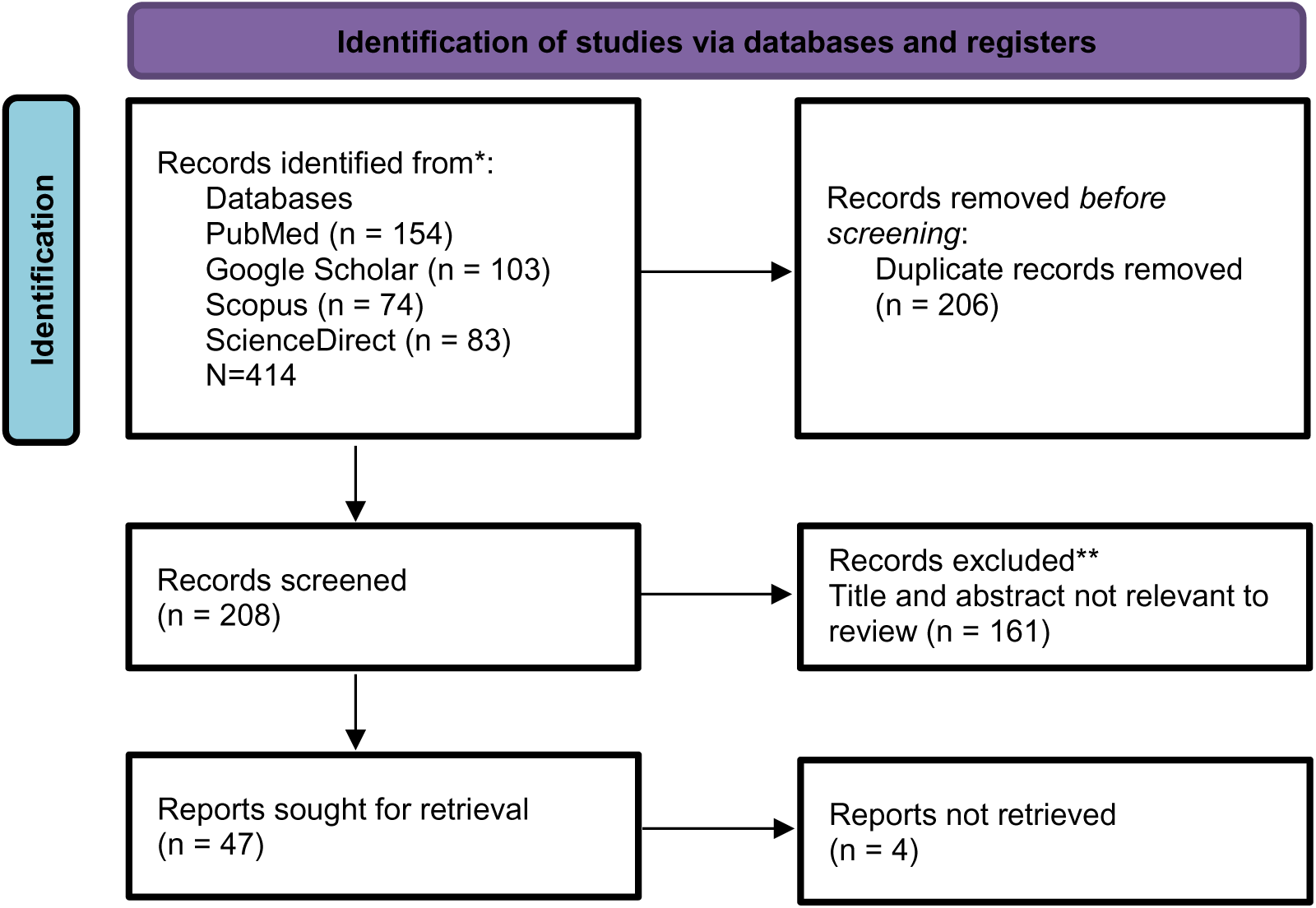
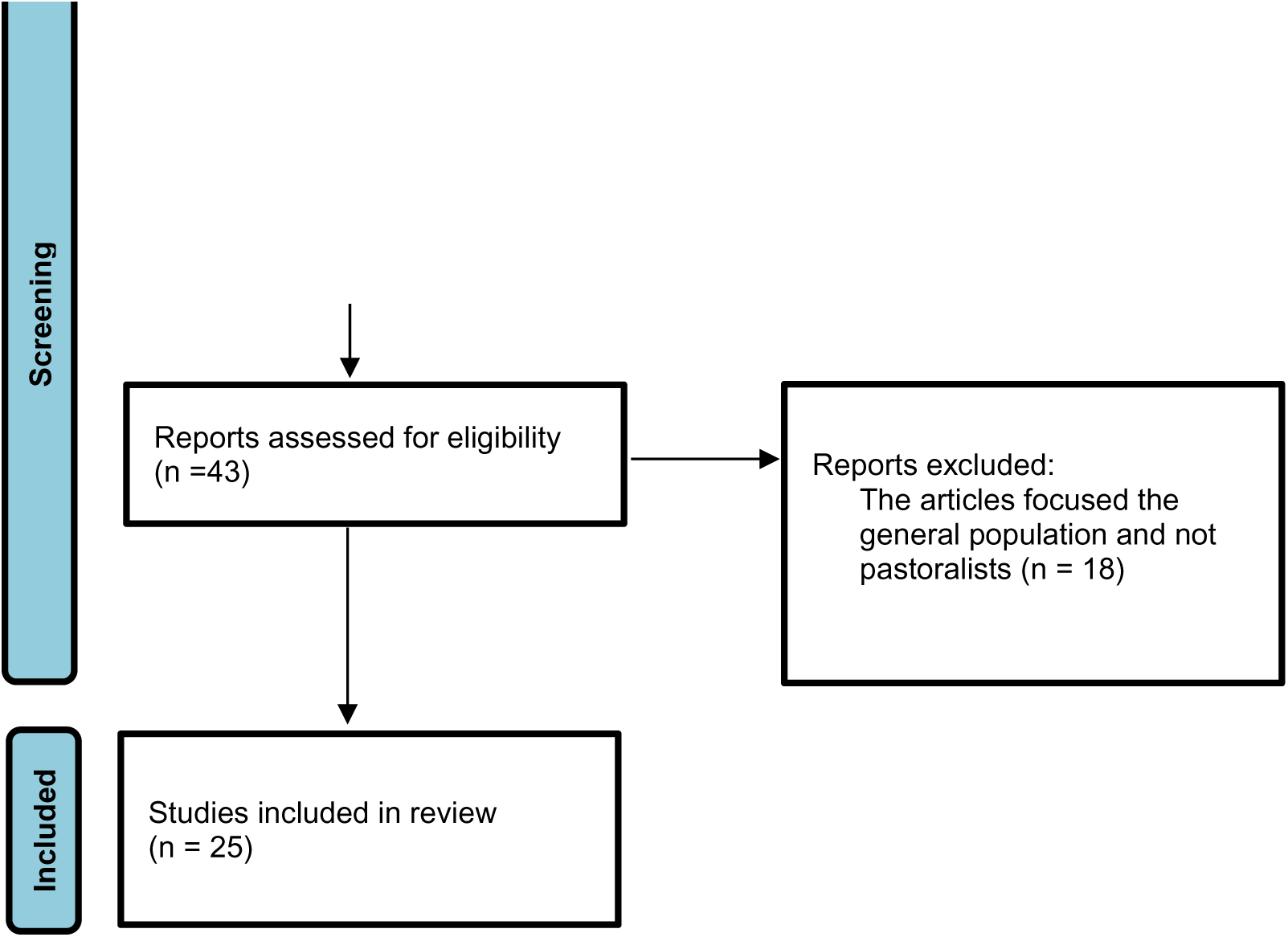
PRISMA Flowchart.

### 2.6 Data Extraction

The extraction in this study is done first by designing data extraction forms. The main information considered in this case is the research aims and objectives, key findings, study settings and the methodology adopted in each study. Before the extraction, the reviewer carried out a pilot test to refine these forms and ensure their consistence with other reviewers. The ration of this pilot testing is to identify any ambiguities in the data extraction process.

### 2.7 Data analysis and synthesis

The extracted data is then synthesised and analysed into common patterns and themes emerging from the existing literature on the issue being investigated. The analysis, in this case, is considered important in providing a comprehensive understanding of the health-seeking behaviours among the nomadic pastoralists based on the existing evidence. The recurring themes and patterns are also vital in the development of this study as they indicate the relevance of findings on the research objectives.

## Chapter Three: Results

### 3.1 Characteristics of the selected studies

The included studies are from diverse areas across LMICs, indicating the depth of evidence used in assessing the determinant of health-seeking behaviours among pastoralists. For instance, out of 25 sources selected, 9 (Ahmed et al., 2018; Ali and Woldearegai, 2019; Henok and Takele, 2017; Hussen et al., 2013; Jackson and Hailemariam, 2016; Kaba and Mariam, 2013; Khogali et al., 2014; Sima et al., 2017; Tessema et al., 2019) were based in Ethiopia. Another country considered in Kenya, in which 6 (Byrne et al., 2016; Caulfield et al., 2016; Diaz, 2017; John et al., 2022; Kenny et al., 2021; Pertet et al., 2018;) sources were based on pastoralist communities in Kenya. Two studies (Barasa and Virhia, 2022; Shayo et al., 2015) focused on pastoralists from Tanzania, Nigeria also had 2 studies (John et al., 2015; Okeibunor et al., 2013) focusing on pastoralist communities. Other countries had one study focusing on the health issues among pastoralist communities. These are (Ahmed et al., 2018) in Mali, (Atekem et al., 2022) in Cameroon, (Duale et al., 2023) in Somalia, (Lechthaler et al., 2018) in Chad, (Lô et al., 2016) in Mauritania and (Seck et al., 2017) in Senegal.

The studies were also based on different methods, which were important in promoting the robustness of the findings or evidence used to meet the research objectives. These included mixed methods research (Ali and Woldearegai, 2019; John et al., 2015; Barasa and Virhia, 2022; Kaba and Mariam, 2013; Lô et al., 2016; Pertet et al., 2018; Sima et al., 2017), qualitative research (Ag-Ahmed et al., 2018; Atekem et al., 2022; Byrne et al, 2016; Caulfield et al., 2016; Duale et al., 2023; Henok and Takele, 2017; Jackson and Hailemariam, 2016; John et al., 2022; Kenny et al. 2021; Okeibunor et al., 2013) and quantitative research studies (Ahmed et al., 2018; Diaz 2017; Hussen et al., 2013; Khogali et al., 2014; Lechthaler et al., 2018; Seck et al., 2017; Shayo et al. 2015; Tessema et al., 2019). Including sources with a wide range of methodologies was vital in complimenting each other in terms of reliability and the quality of information gathered.

**Table 2:**
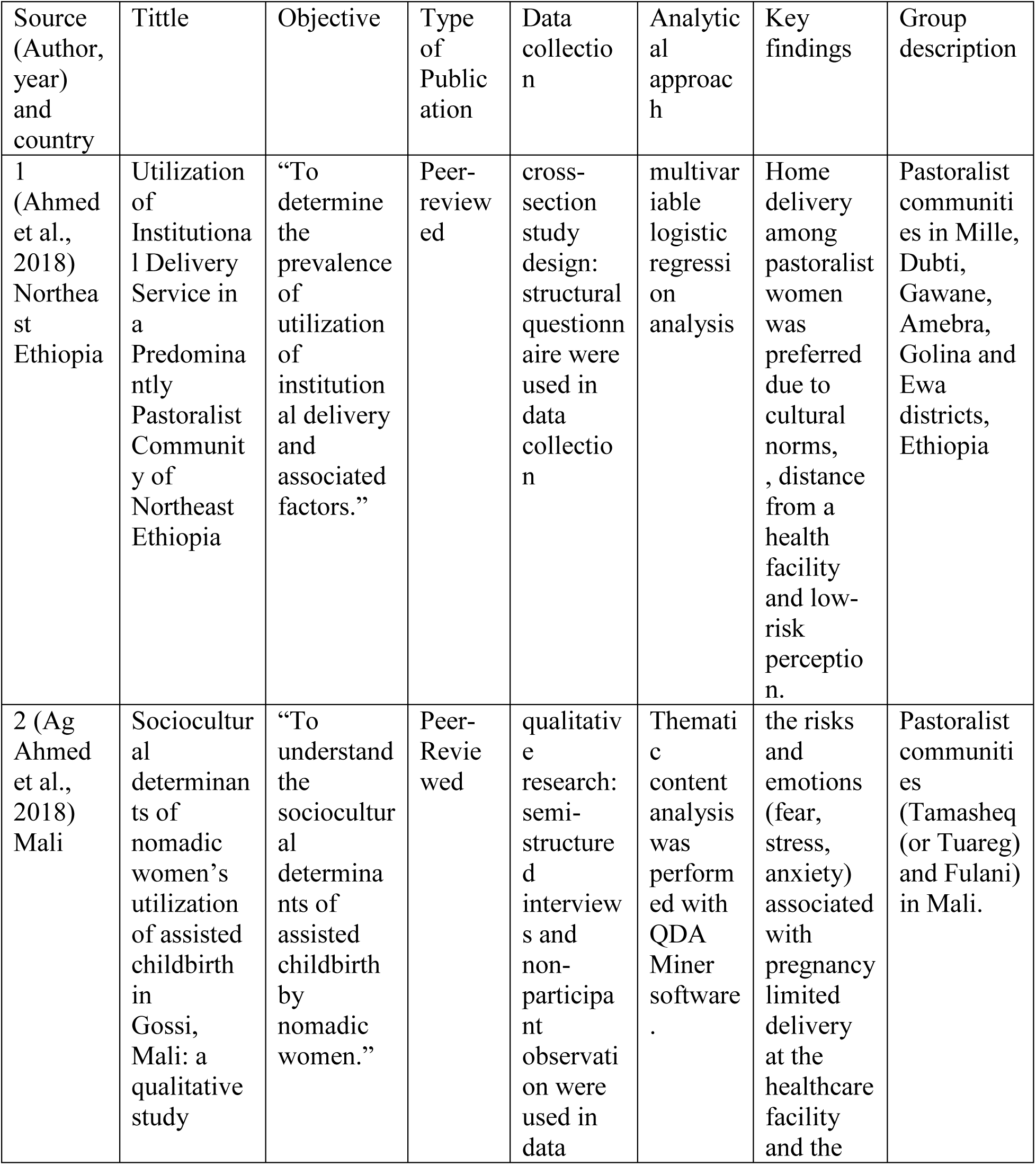
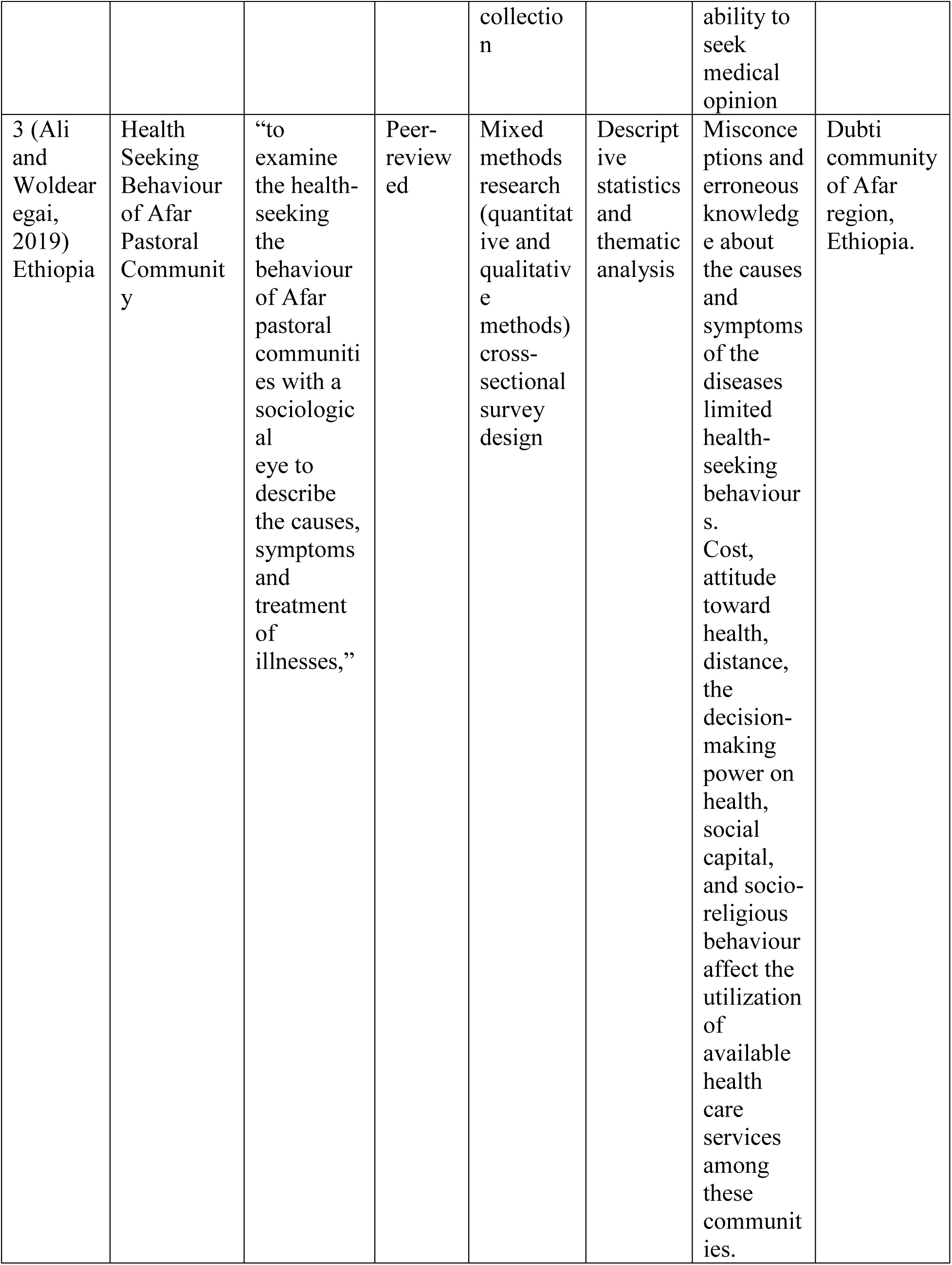
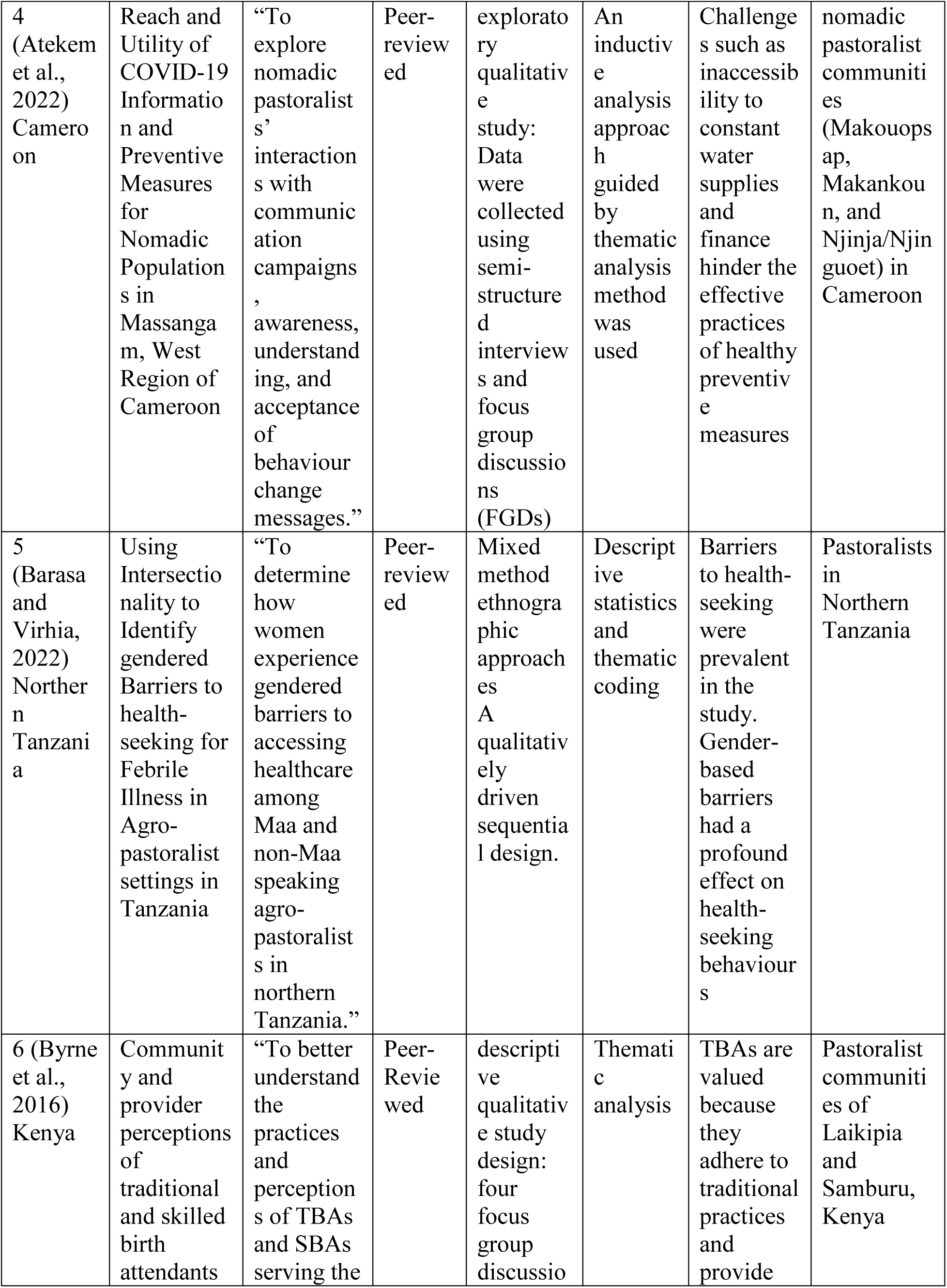
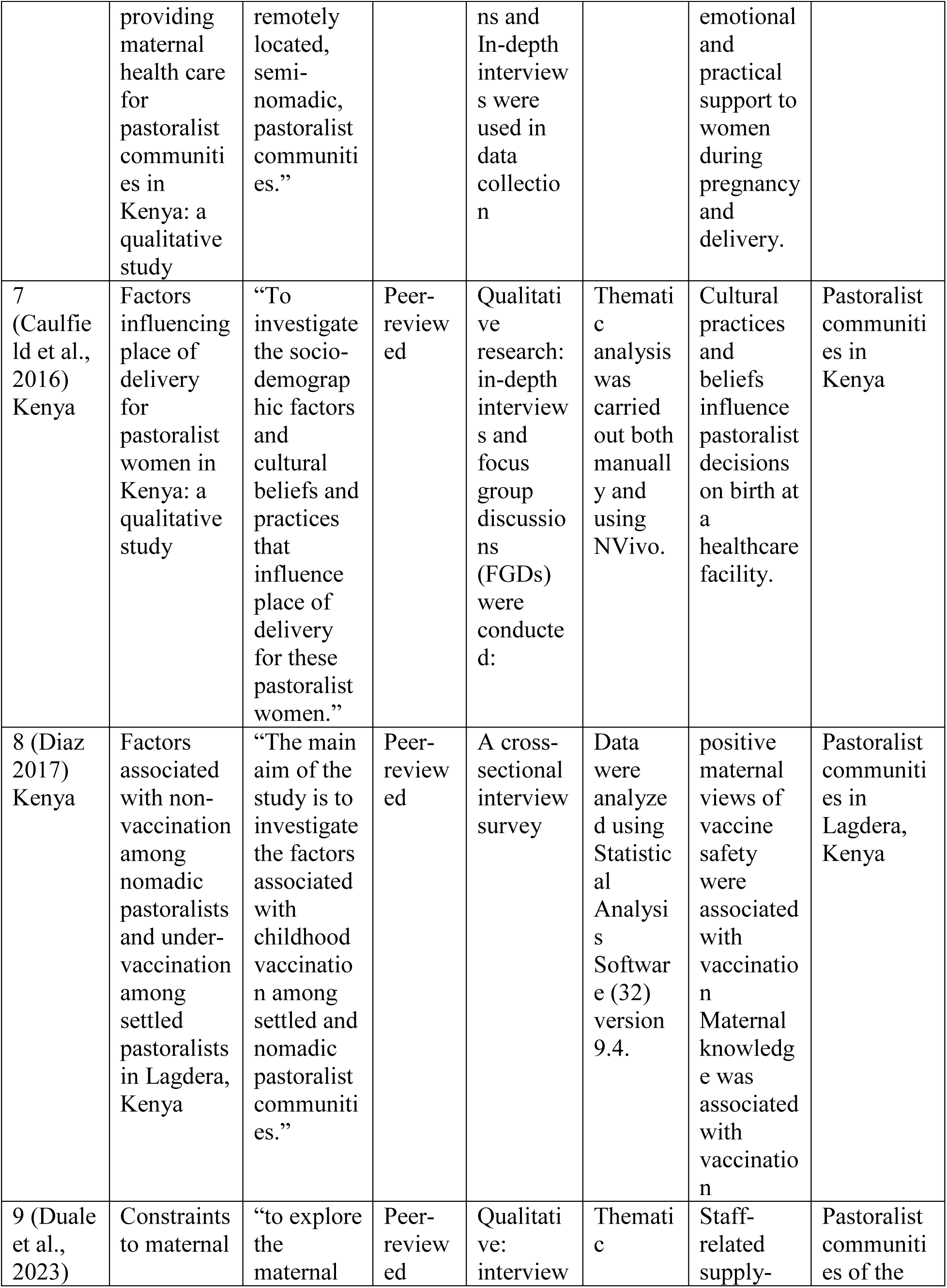
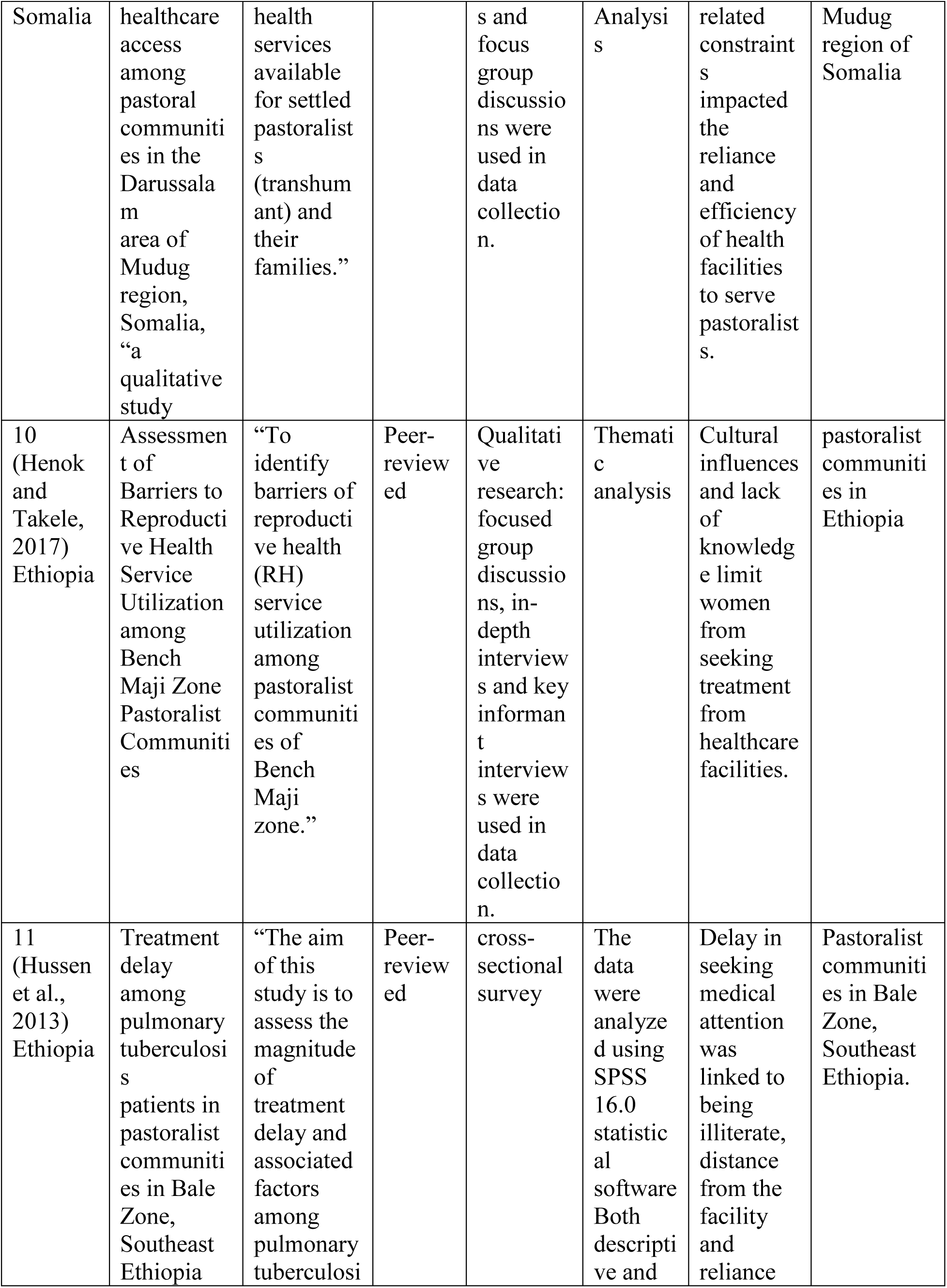
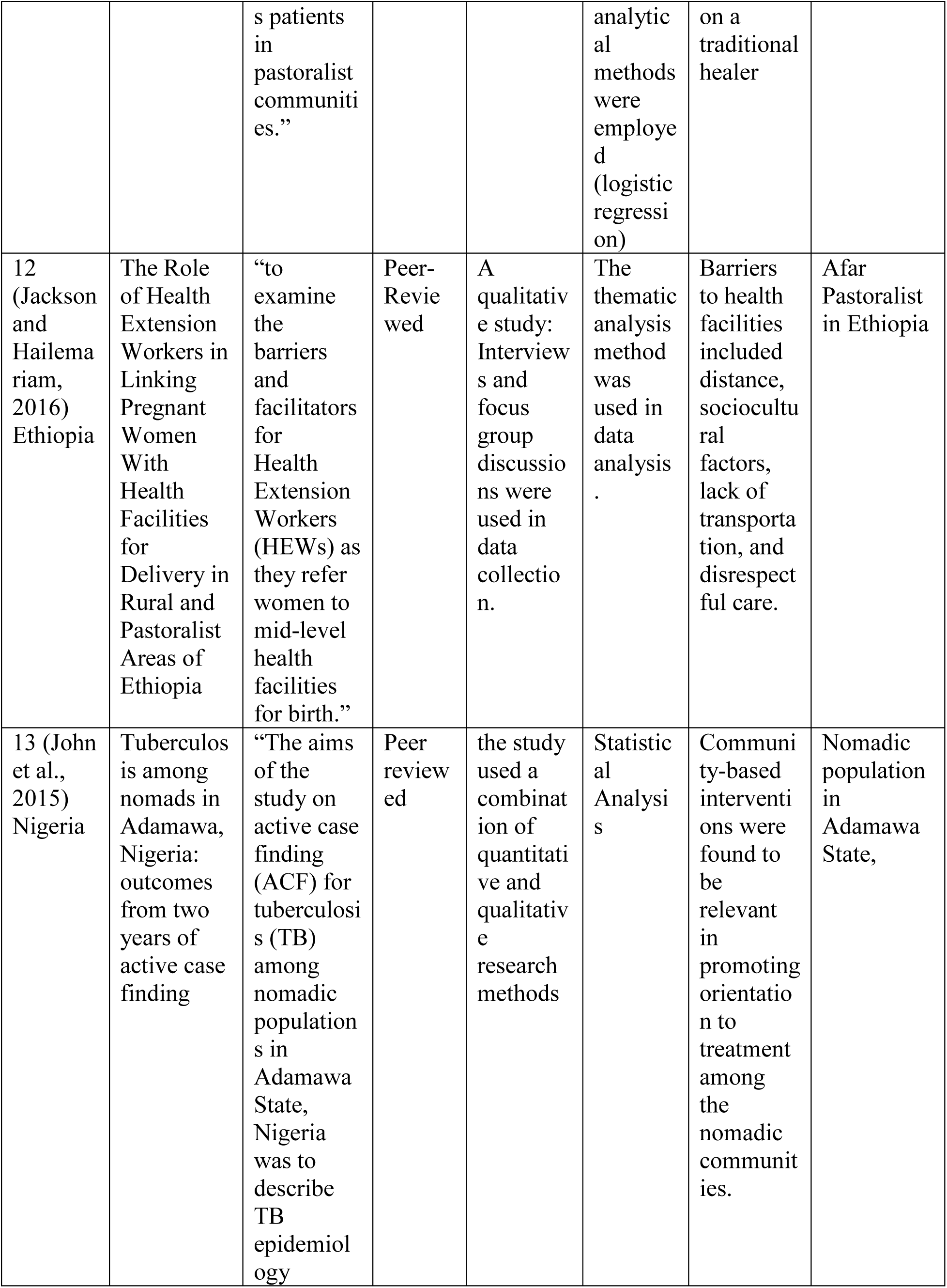
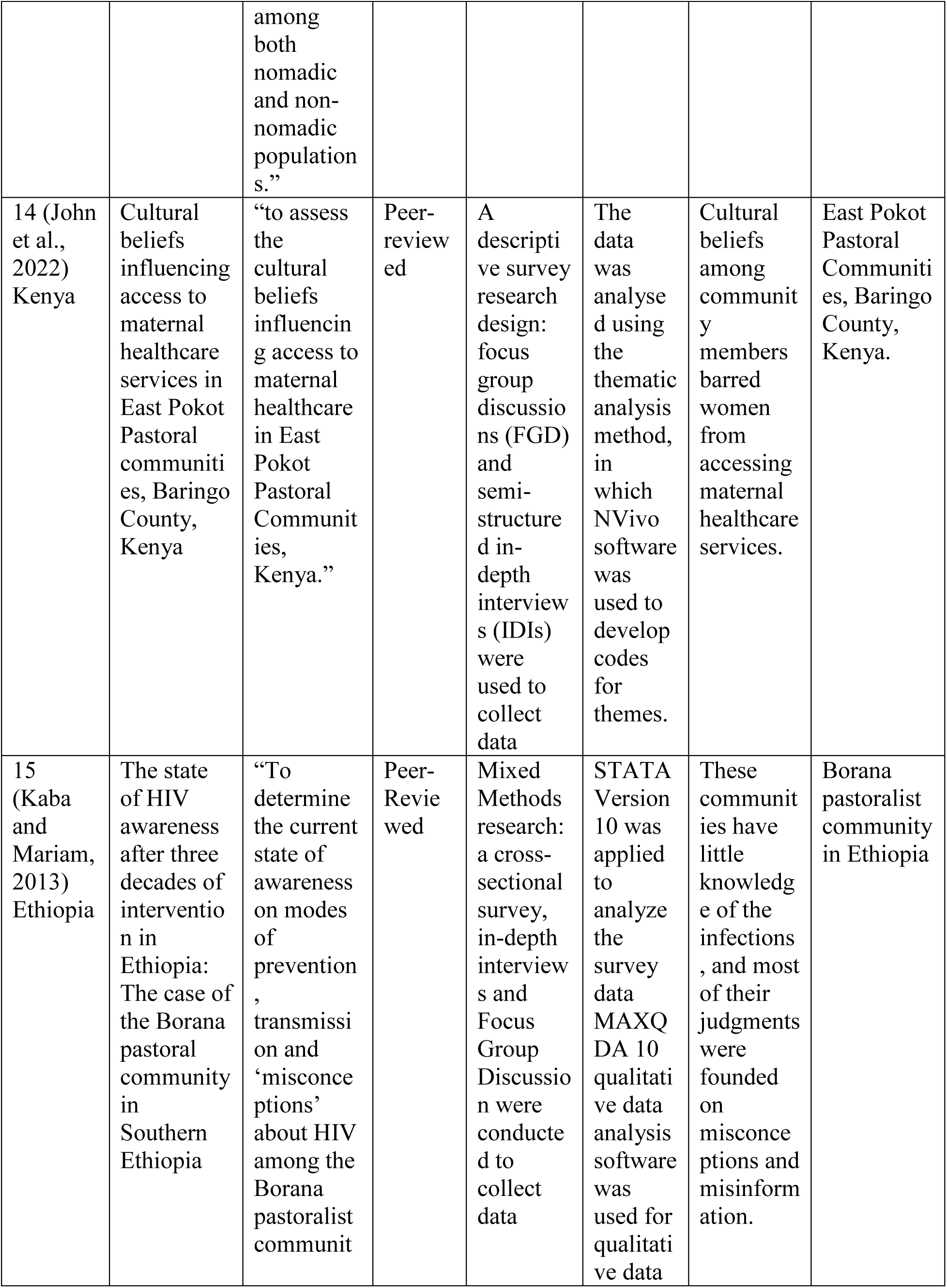
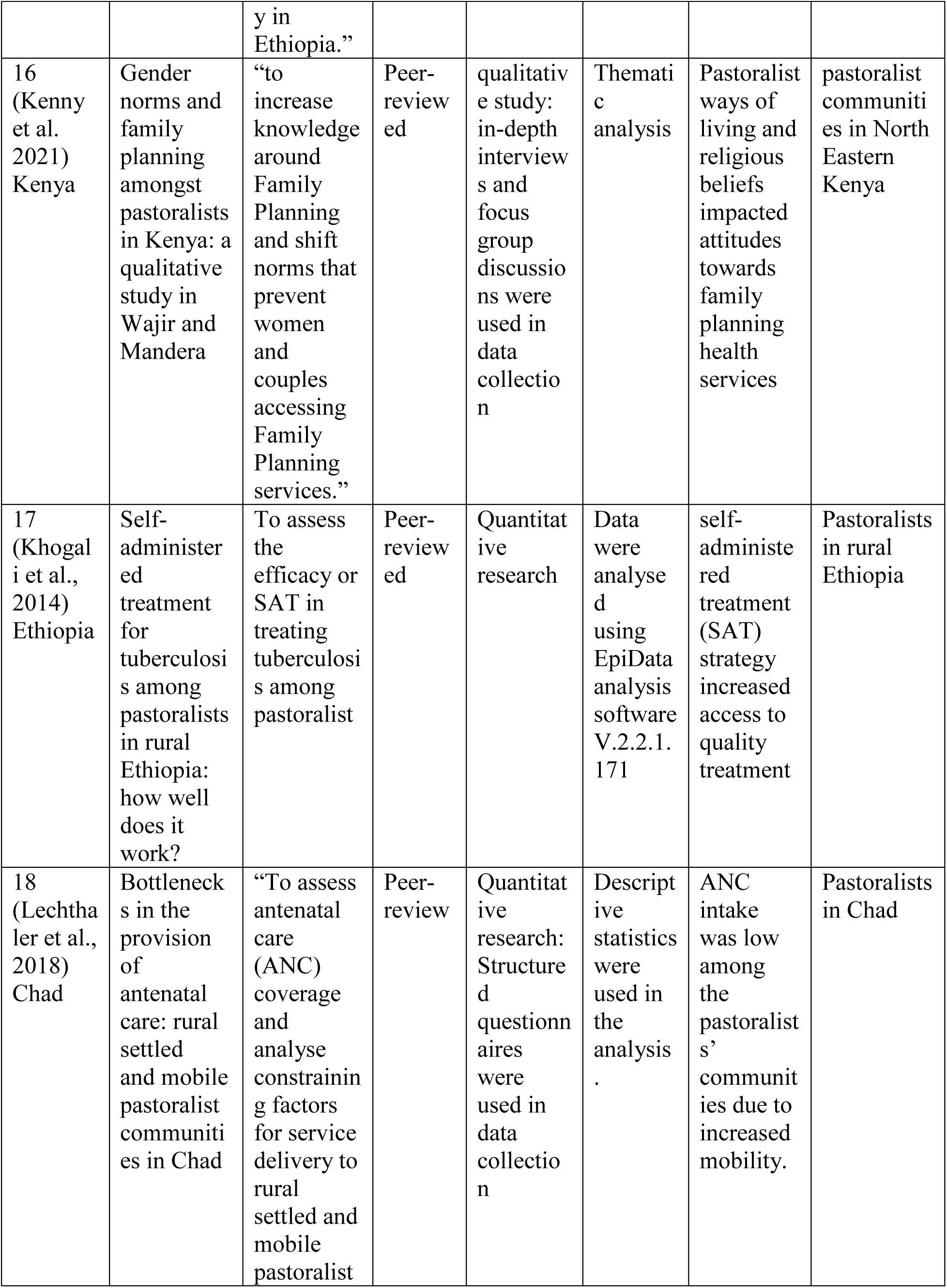
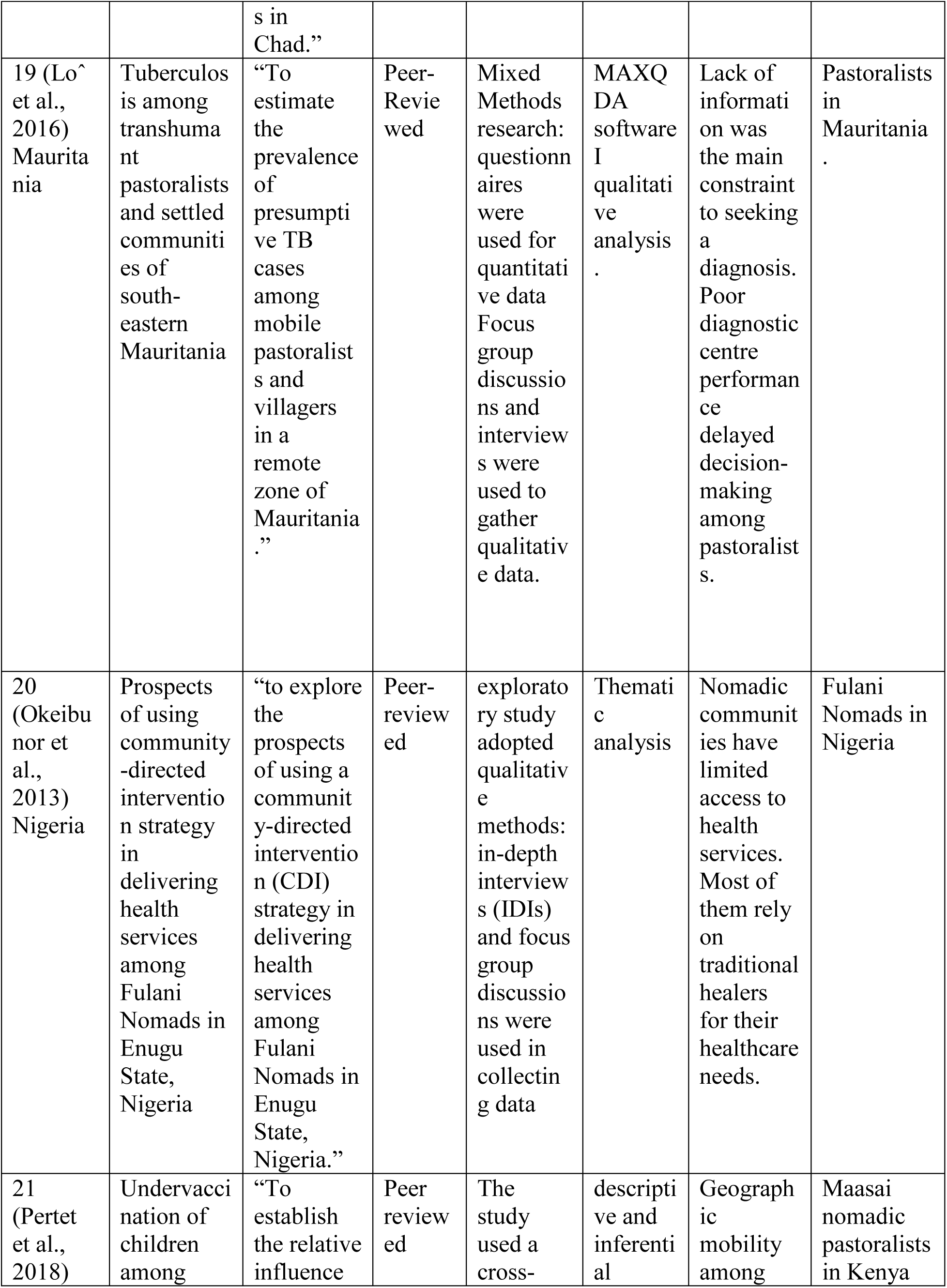
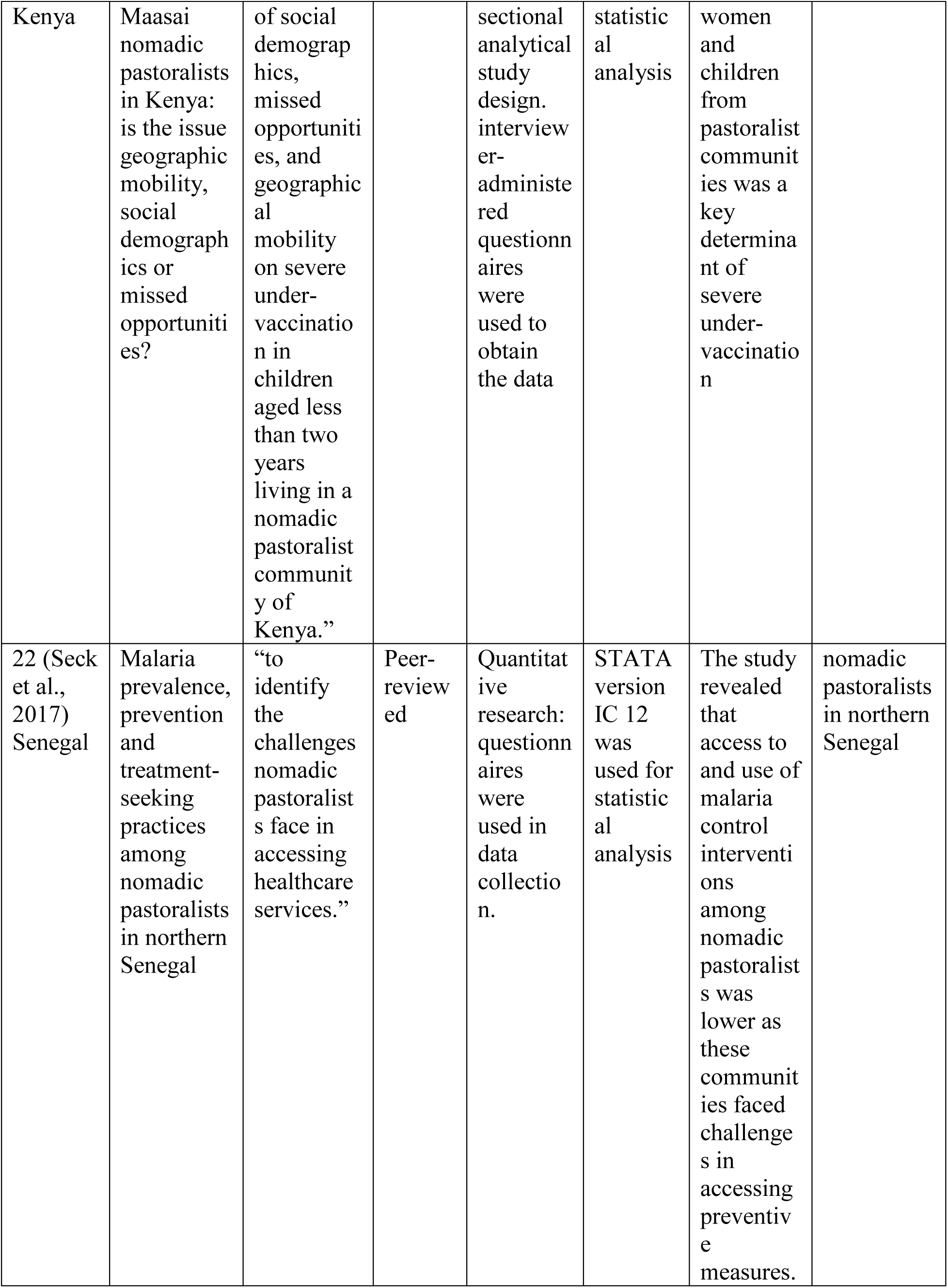
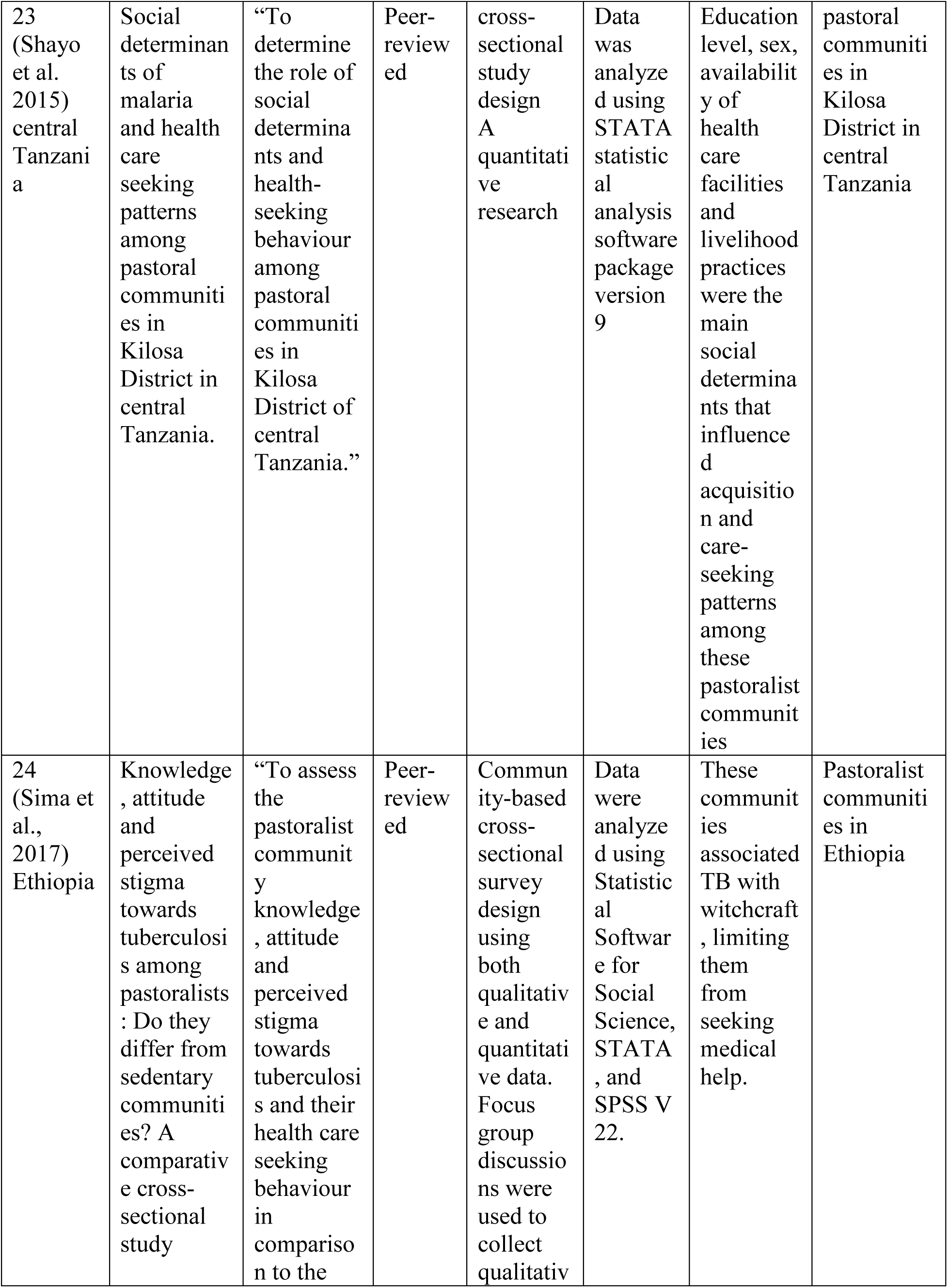
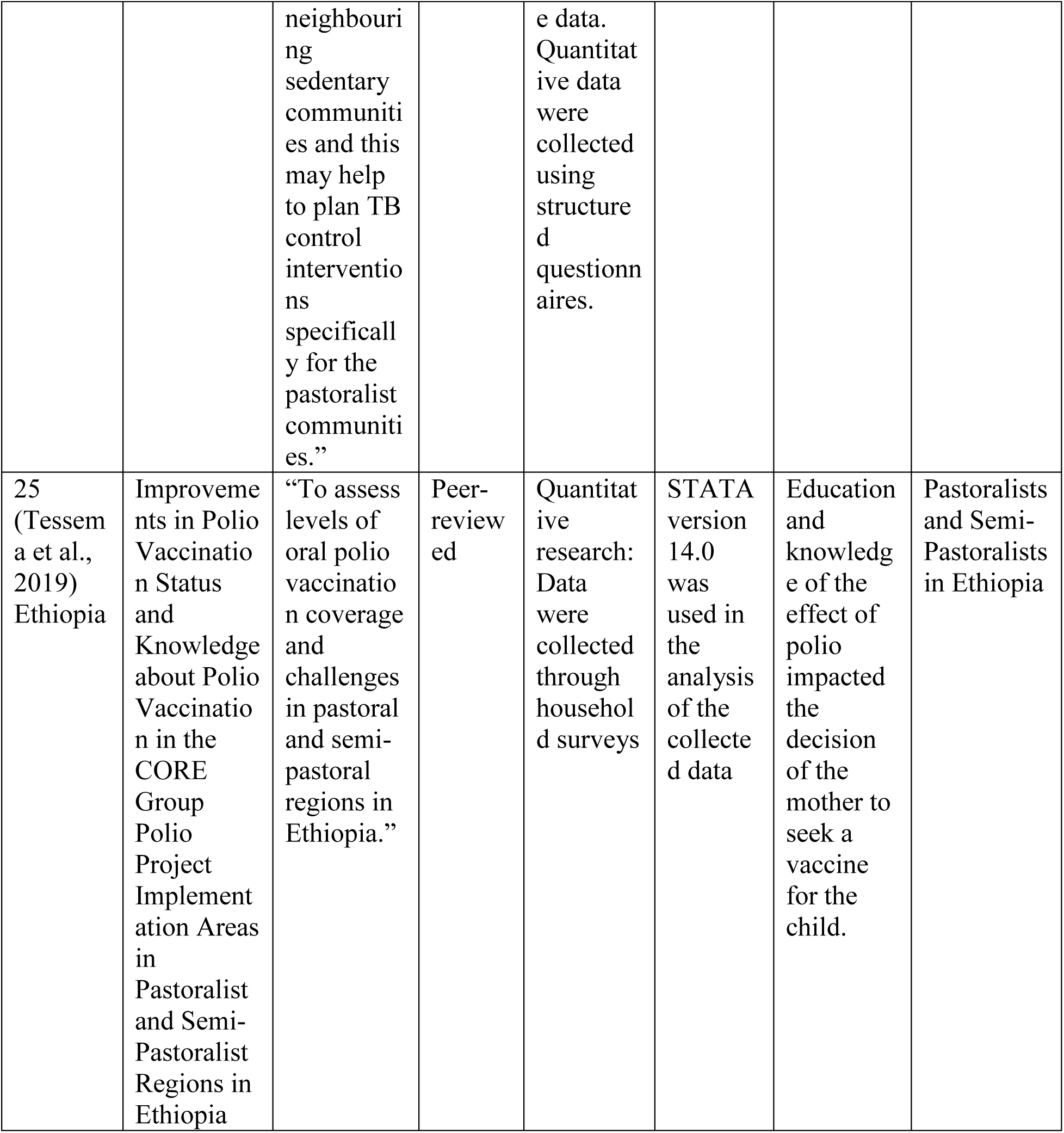
Selected studies.

**Table 3:**
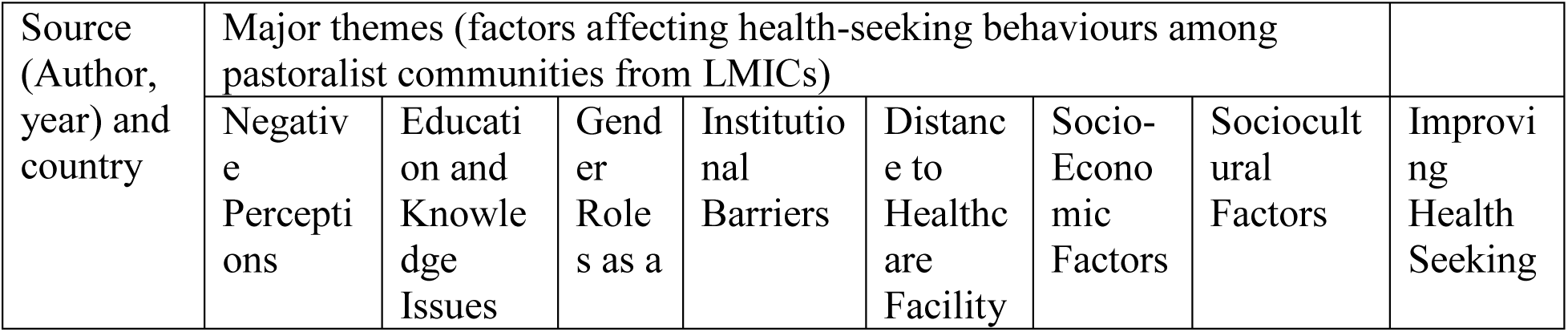
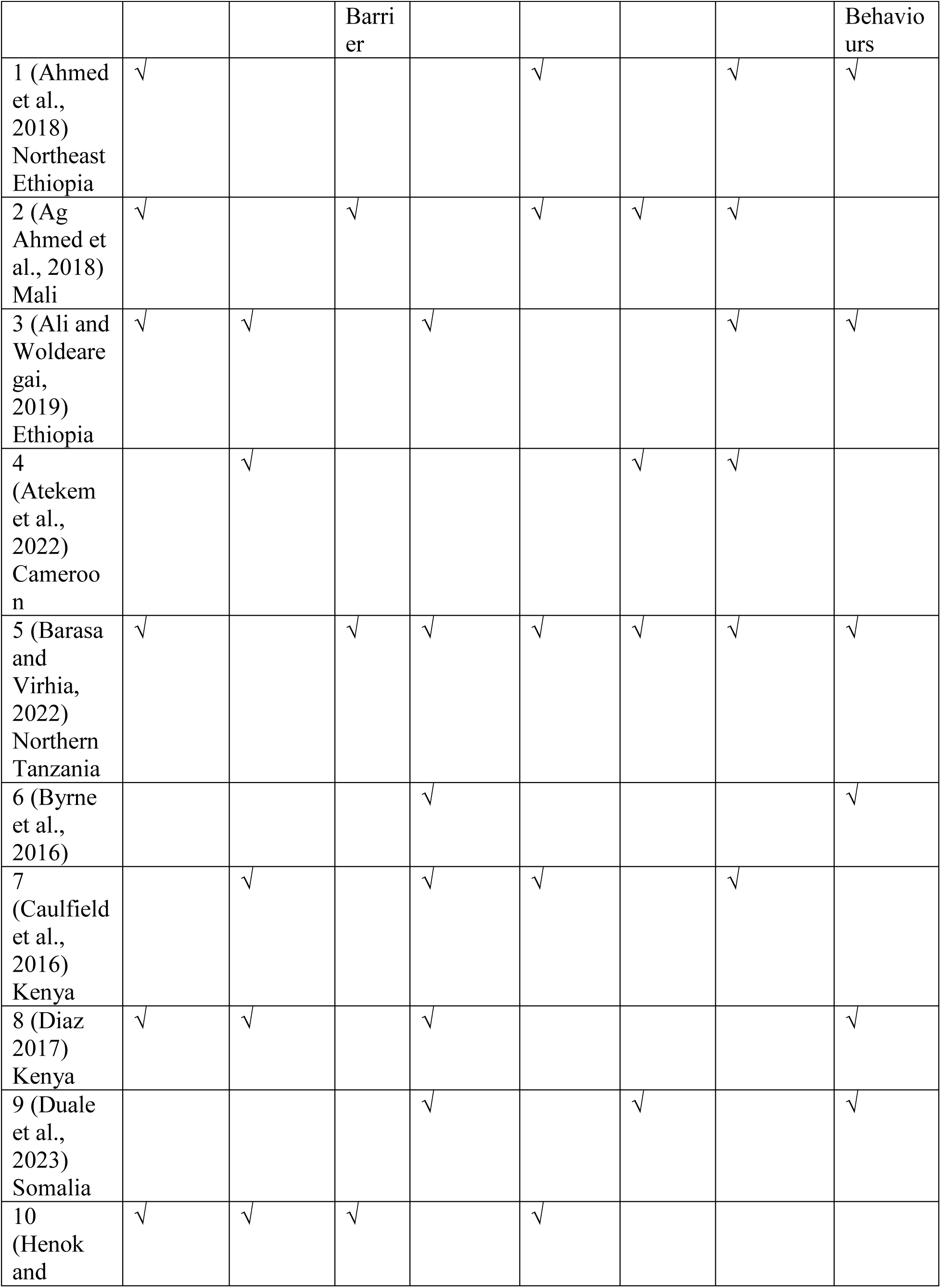
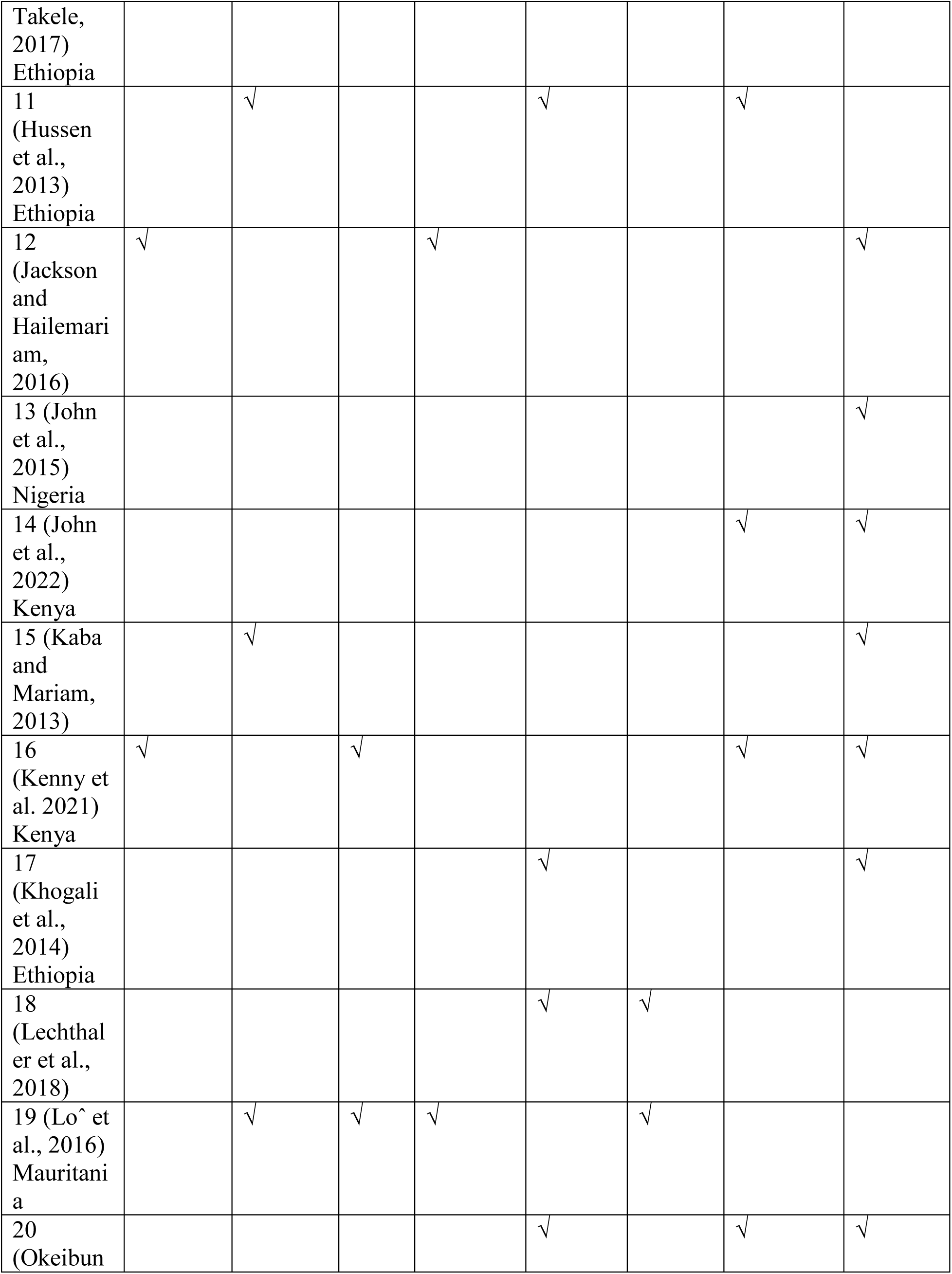
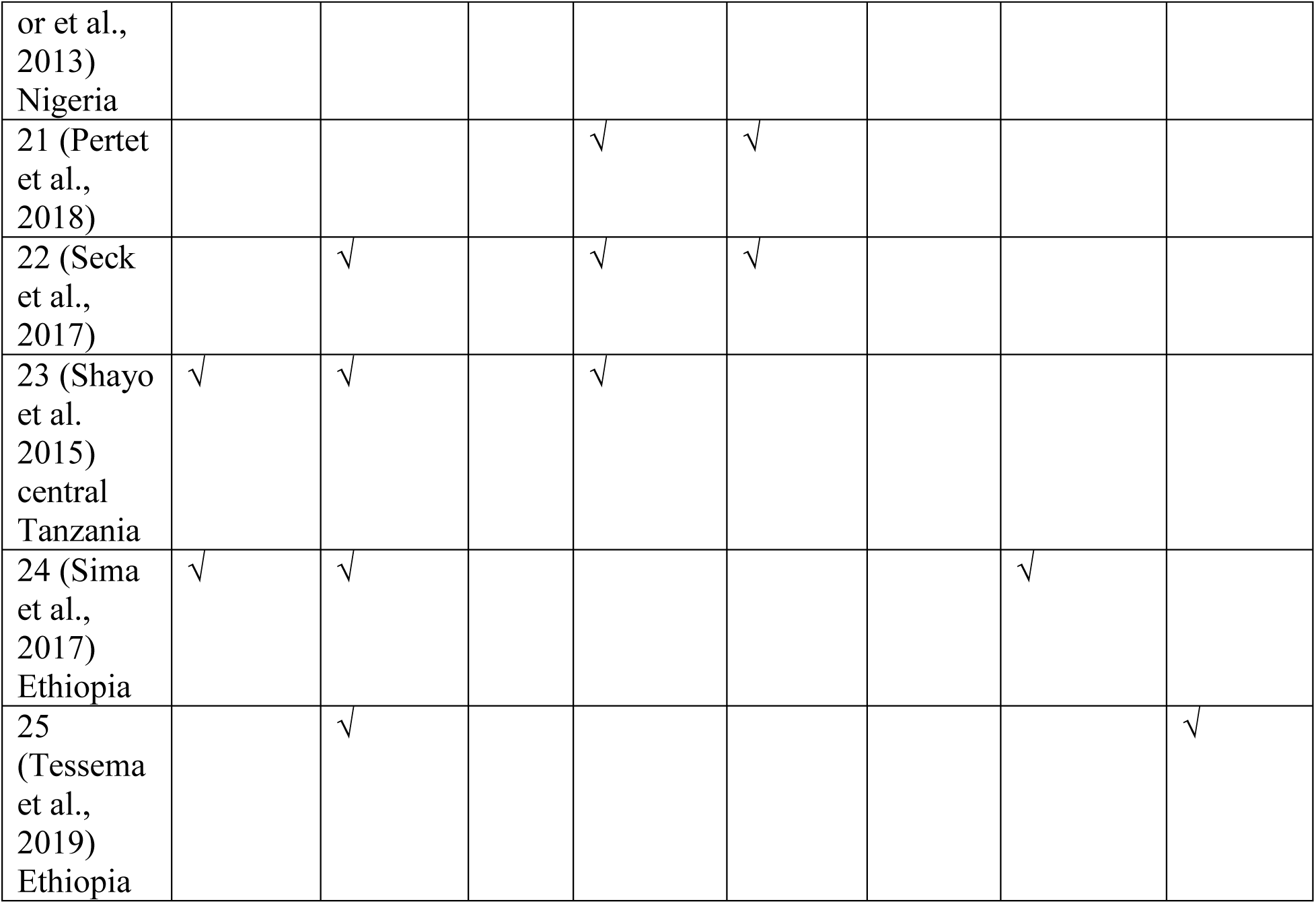
Major Themes identified from the selected studies.

### 3.2 Negative Perceptions

One of the issues impacting the health-seeking behaviours among these communities included their views on the illness or the symptoms involved. Barasa and Virhia (2022), in this case, found that the seriousness of the disease would be considered when the symptoms affect their activity of daily living. In that regard, when the symptoms are not viewed as severe, they would not alert an individual to seek any medical attention from a qualified physician. Barasa and Virhia (2022) noted the misconception about some symptoms, especially in the case of fever, noting that it was something to be anticipated and lived with. According to Ag Ahmed et al. (2018), nomadic women in Gossi regarded childbirth as a natural process that did not necessitate medical intervention, thus preferring not to seek any medical assistance. Ali and Woldearegai (2019) also noted that these people used misconceptions and perceived causes of the illness, such as improper eating habits, curses, sins, witchcraft, and environmental conditions. This led to complexity in their illness, minimizing their motivation to visit healthcare facilities. According to Sima et al. (2017), pastoralists in Ethiopia had negative attitudes towards diseases such as TB, including feelings of fear or embarrassment they associated with the disease and a higher overall perceived stigma linked with TB. Such attitudes have been traced to knowledge gaps and a lower level of understanding of the dynamics around the disease.

In some aspects, the perception also limited these people from adopting measures that could have been vital in promoting well-being and preventing adverse health issues. For instance, Shayo et al. (2015) noted the concept of negative perception not only on the causes of the disease but also on the interventions promoted to fight diseases such as malaria as a significant determinant of low health-seeking behaviours. In support of this, Kenny et al. (2021) indicated that pastoralists in Kenya were reluctant to seek family planning services as they had negative attitudes towards these services, especially in the case of modern family planning (FP) methods used to prevent serious negative health outcomes. In the same case, Ahmed et al. (2018) noted that women from pastoralist communities were reluctant to seek treatment from healthcare facilities, especially during delivery, based on the fear of being attended by a male clinician. Henok and Takele (2017) also found similar concerns, noting that most health professionals and health extension workers in these communities are predominantly male, and women do not see them as suitable to assist them during the delivery. According to Jackson and Hailemariam (2016), this issue makes most women fail to seek medicating services, noting that the majority of women from pastoralist communities preferred giving birth attended by TBAs or at home supported by family members or neighbours rather than giving birth in health institutions.

The issues of health-seeking behaviours among the pastoralists were also linked with the value they played on their animals, noting that seeking health care services may limit their ability and time to take care of the animal. In this case, Barasa and Virhia (2022) state that it is a common aspect for pastoralist communities to consider their herding activities over their health, noting that it would affect their reasonability in taking care of the animals.

As Barasa and Virhia (2022) associated barriers to positive health-seeking behaviours with poor interpretations, misconceptions, and perceptions of the symptoms, other scholars indicate that the entire issue occurs due to communication barriers. For instance, Ali and Woldearegai (2019) indicate that communication challenges between health professionals and patients, especially those from pastoralist communities, limit their understanding of the issues while also lacking the capacity to communicate such issues to their healthcare providers effectively. The issues of individual views were also promoted in a study by Diaz (2017), who noted that when women have positive views on the importance of vaccination in promoting the health of their children, they are inclined to seek medication attention, especially through securing vaccinations for their children. In this regard, when there is enough knowledge of the importance of vaccination, women from pastoralist communities tend to take their children for vaccination.

### 3.3 Education and Knowledge Issues

According to Ali and Woldearegai (2019), the negative perception and conceptualization of diseases (which further shapes health-seeking behaviours) have been subject to erroneous knowledge and misconceptions about the causes and symptoms of illnesses. This case was common, especially among community members or pastoralists with no or low education levels (Ali and Woldearegai, 2019). According to Hussen et al. (2013), illiteracy and low levels of education led to a failure to understand the symptomatic issues, which later led to a prolonged delay in seeking medical attention among pastoralists. Sima et al. (2017) noted that the pastoralist community portrayed a significant knowledge gap about the signs and symptoms, causes, mode of transmission, treatment and prevention of TB. Such knowledge affected them as they failed to seek medical attention. Furthermore, Kaba and Mariam (2013) noted that low knowledge of the nature of the disease and its symptoms increased misconceptions about the transmission and preventive measures that people can adopt. Shayo et al. (2015) also indicate that most of the pastoralists had inadequate knowledge of diseases such as Malaria, which was an important factor in limiting their intentions to seek medication or medical advice. This was also a common issue among the non-educated pastoralists, indicating the importance of knowledge and education on health-seeking behaviours. Seck et al. (2017) also raised this issue, indicating that most nomadic pastoralists have no knowledge of the causes and symptoms of serious diseases such as malaria, limiting them from seeking medical attention.

The concept of knowledge about the disease was also a common factor in the study by Atekem et al. (2022), who indicated that the nomadic people had limited knowledge of COVID-19 symptoms; therefore, they were reluctant to adapt to any preventive measures or seek medical attention. Atekem et al. (2022) noted that such unawareness was a significant health threat because most of these people were also not aware of the transmission routes for various infections. Diaz (2017), who assessed the issue of knowledge among mothers in the pastoralist communities in Kenya, noted that adequate knowledge is a significant factor in changing the attitudes of these women, making them prefer to vaccinate their children. The change in attitude is based on the knowledge these women have regarding the importance of vaccines on the overall health and wellbeing of the children. Henok and Takele (2017) further identified that poor decisions that women from pastoralist communities make are majorly due to a lack of knowledge on better planning and delivery aspects and other issues related to their health. Tessema et al. (2019) also indicated that maternal education and awareness about the threats of polio and the impact of vaccines would make people diligent in health matters, increasing their intentions to seek these vaccines. According to Caulfield et al. (2016), lack of awareness and education regarding the risks of delivering at home plays a significant role in pastoralist women’s decision to deliver at home. Lô et al. (2016) indicate that although information and public health campaigns may be provided, they become useful for the general population because they are done for a limited time and fail to get to the pastoralist communities who are always on the move.

### 3.4 Gender Roles as a Barrier

Another important outcome in this review is that gender role played an important role in limiting people from seeking medical attention. In their assessment, Barasa and Virhia (2022) found that young men and those bellow the age of 40 in Tanzania were considered the primary herders among the pastoralist communities, and therefore, they would not be required to neglect their role. In particular, leaving the livestock to focus on their health would be considered neglect to their primary role of taking care of the herds (Barasa and Virhia, 2022). Lô et al. (2016) indicated that seeking a diagnosis would be considered an activity that would make these people out of work (herding), thus preferring to work rather than visiting healthcare facilities for healthcare issues.

Apart from gender roles, the power difference between different genders within the pastoralist communities played a vital role in impacting the ability to seek health services or treatment. Kenny et al. (2021) in this case noted that Men were often the decision-makers concerning family planning services, and the desires of women for smaller families were often outweighed; as such, these women would seek family planning services upon being approved by their husbands. Ag-Ahmed et al. (2018) also note that power dynamics and gender roles played a role in the decisions of nomadic women to use or not use assisted delivery. In particular, women from these communities often had controlled decision-making power in their families as they were expected to follow the desires of their husbands. Ag-Ahmed et al. (2018) further found that women from pastoralist communities faced resistance from their husbands who did not want them to give birth in a healthcare centre. Henok and Takele (2017), in this case, indicated that most husbands do not see it fit for their wives to be attended by male professionals; thus, they disapprove of any healthcare services that may bring such an encounter.

### 3.5 Institutional Barriers

The issue of the health system barrier has been another critical concern affecting the health-seeking behaviours among the nomad pastoralists. In this case, Barasa and Virhia (2022) indicate that these people do not depend on a regulated framework of care as they consistently rely on self-administered allopathic treatment and self-treatment with herbal medicine, indicating the lack of medication and poor staff attitudes toward these communities. Duale et al. (2023) also found institutional barriers such as staff-related constraints, lack of necessary training for crucial areas, capacity of the staff, and limitations in infrastructure and resources as vital challenges limiting their access and utilization of health services. Additionally, views of medication effectiveness and treatment costs are significant concerns, making these people avoid engaging with health facilities (Barasa and Virhia, 2022). Similarly to Ali and Woldearegai (2019), these communities were also making choices based on the availability of required medicine, attitude towards health care services and the overall cost of health care services. Jackson and Hailemariam (2016) also, in assessing the relevance of healthcare staff in promoting health-seeking behaviours, noted that the fear and encounters of disrespect and abuse by healthcare professionals impacted the willingness of women from pastoralist communities to seek facility delivery. Lô et al. (2016) also noted that disrespectful treatment from health workers and the perceived low quality of service in the Mauritanian healthcare system made pastoralists not visit these facilities for any healthcare need. Caulfield et al. (2016) identified similar issues among pastoralists in Kenya, noting that the perception of unfriendly and disrespectful treatment care at health facilities differed from the warm and caring experience provided by TBAs, thus making women from these communities not seek medical opinion or services from the healthcare facility.

The concept of “limited resources” is a significant determinant because, irrespective of how informed these people are, the lack of relevant facilities would still limit their access to healthcare services (Duale et al., 2023). Diaz (2017), in this case, indicates that irrespective of the willingness of women from the pastoralist community to improve the health status of their children, the obstacles they face in securing childhood vaccinations limit their intentions to improve the health status of their children. Diaz (2017) further notes that this has been a significant problem among pastoralists in Kenya because there are no tailored vaccination programmes that would promote vaccination practices for both nomadic and settled pastoralist communities. Pertet et al. (2018), who also assessed vaccinations among the children of pastoralists in Kenya, noted that vaccine stock outs were a key factor limiting access to better health services among these communities. Accessibility issues were raised in the study by Seck et al. (2017), who indicated that nomadic pastoralists have not been accessing preventive equipment such as mosquito nets, thus limiting them from utilising malaria control interventions.

The issues of visiting healthcare institutions or facilities have also been impacted by how individuals make decisions about their care plans. According to Barasa and Virhia (2022), people make decisions about health care services based on their perceptions of the capacity of health facilities to cure their condition or help them manage their health concerns. In particular, when they experience a health system that falls short of their expectations, they end up considering self-treatment as a means of saving both time and costs associated with going to the hospital (Barasa and Virhia, 2022). Similar concerns were raised by Ali and Woldearegai (2019), who indicated that the choice of healthcare facility and the decision to be involved in one was influenced by individual briefs on the services offered and the trust in the effectiveness of medication. Shayo et al. (2015), in this case, indicated that most of the pastoralists had a negative perception of the effectiveness of mosquito nets in protecting them from malaria, thus limiting them from the available intervention. However, according to Byrne et al. (2016), the issue is how the services provided resonate with the needs of the people involved. For instance, pastoralist communities in Kenya were found to prefer TBAs over skilled birth assistants because they believed that TBAs adhered to traditional practices while providing practical support to these women during pregnancy and delivery (Byrne et al., 2016). Jackson and Hailemariam (2016), in the same case, noted that some prefer interacting with Health Extension Workers (HEWs) as they trust them in making decisions about the place for delivery and medical needs. This indicated the importance of an integrated model in solving health issues among pastoralists.

### 3.6 Distance to Healthcare Facility

Another important institutional barrier is the increased distance to the health facilities. According to Barasa and Virhia (2022), lack of access to formal healthcare facilities due to movements and distance from the facility has been limiting people from the pastoralist communities to seek medical attention. The same issues emerged as a significant concern among the pastoralists in Ethiopia as they attributed the prolonged delay in seeking treatment for pulmonary tuberculosis to distance to the healthcare facility (Hussen et al., 2013). These authors indicated that most of these patients live more than 10 kilometres from the dearest healthcare facility, making them reluctant to visit for regular checkups and medication. In the assessment of the nomadic people in Nigeria, Okeibunor et al. (2013) indicated that the constant movement of livestock creates an artificial barrier, giving these communities limited access to health services. Ahmed et al. (2018) noted that the distance would continue increasing the gap in accessibility of healthcare services because of the seasonal mobility among the pastoralists’ communities. In the same case, Ag-Ahmed et al. (2018) travel long distances to seek medical attention, making pastoralists prefer alternative methods for addressing their issues, such as relying on TBAs.

Caulfield et al. (2016), who assessed the choice of women on where they prefer to give birth, noted that pastoralist women were substantially influenced by factors such as poor roads, distance and limited transport, making them prefer giving birth at home or not seek any maternal help from the healthcare facility. Seck et al. (2017) indicate that nomadic pastoralists frequently travel in remote areas in addition to their infrequent contact with health systems, which poses challenges in accessing healthcare services. These issues limit access to health services, resulting in delayed care-seeking, particularly for asymptomatic infections, which contribute to ongoing transmission (Seck et al., 2017). In exploring this issue further, Lechthaler et al. (2018) indicated that irrespective of women having adequate information about ANC visits, the mobility nature of these communities made them not adequately use such services for their maternal health concerns. In terms of distance from the healthcare facility, Pertet et al. (2018) indicated that the geographic mobility of women and children from nomadic pastoralists has been a key determinant of severe under-vaccination in Kenya. These children would, therefore, miss the opportunity to be vaccinated as they are usually away from the health facilities. Khogali et al. (2014) also cite the issues of distance and mobility factors as these communities are in constant movement, failing to adhere to some medications, especially in the case of patients with tuberculosis. Henok and Takele (2017) indicated that these communities are significantly isolated due to long distances to facilities and poor infrastructure. Such low access leads to preferences for poor health practices, such as relying on unsafe delivery methods.

### 3.7 Socio-Economic Factors

The financial ability of the family involved in the pastoralist communities was considered an important determinant of their ability to seek medical attention. Barasa and Virhia (2022), in this case, note that community and household abilities shape access to medical resources, including medical charges money for buying over-the-counter medication. Duale et al. (2023) further appraise the significance of financial constraints, indicating that since pastoralists are predominantly poor people who are unable to pay an expensive user fee for blood transfusions, C-sections, and similar maternal health services, they are reluctant to seek medical attention from healthcare facilities. Lechthaler et al. (2018) also noted that in addition to the acceptability of modern healthcare services, the affordability aspect increased the barriers to accessibility of these services among the pastoralist communities. Another study based in Cameroon also indicated that finance issues limited the ability and the capacity of community members to adopt effective healthy practices, especially in preventing the pandemic (Atekem et al., 2022). Furthermore, Ag-Ahmed et al. (2018), who assessed the case of pastoralists in Mali, noted that the thought of paying for transportation to healthcare facilities and other medical charges made these people prefer other methods. Lô et al. (2016) also raise these issues, indicating that the financial sides of coping costs for seeking diagnosis among the pastoralists, such as long travel time, are significant obstacles to visiting a TB diagnostic centre to seek medical attention.

### 3.8 Sociocultural Factors

The intention to seek healthcare services among these pastoralists has also been impacted by other issues cultural and social issues. These issues include religious and tribal discrimination in health organisations (Barasa and Virhia, 2022). Kenny et al. (2021) also mention the issues of religious beliefs by indicating that religious beliefs and pastoralist ways of life were limiting factors to women seeking family planning as significant healthcare services among the nomadic and semi-nomadic pastoralist communities in Kenya. Ag Ahmed et al. (2018) in this case noted that traditional beliefs and practices had been significantly impacting treatment-seeking among the pastoralists’ women, especially in Gossi, who believed that giving birth in a healthcare facility would bring bad luck or harm to the child. Caulfield et al. (2016), who also assessed women from pastoralist communities, found that local cultural values associated with women and childbirth play a substantial role in the decision to deliver at home. Furthermore, the beliefs of these women that traditional birth attendants (TBAs) were more experienced and skilled than health professionals made them not seek any medical attention (Ag Ahmed et al., 2018). (John et al. (2022), who also assessed pastoralists in Kenya, noted that cultural beliefs had a prominent role in determining access to maternal healthcare services. The opinion of these communities was also shaped by religious and traditional beliefs held in the communities on pregnancy and giving birth, noting that using maternal healthcare services has a negative effect on women (John et al., 2022). Such factors isolate women from facility delivery, which is safe and crucial for their children.

Additionally, stigmatization faced by women from the pastoralists’ communities by communities limited their intentions to seek or use modern FP methods, irrespective of the impacts of these services on their health (Kenny et al., 2021). According to Kenny et al. (2021), traditional means took precedence, making it seem inappropriate to seek medical attention outside of social and cultural beliefs. Sima et al. (2017) stress the issue of reliance on tradition by indicating that the pastoralist community demonstrated more frequent visits to traditional healers for their medical needs. Ahmed et al. (2018) also noted that the influence of culture, traditional practices and beliefs shaped the use of healthcare facilities among women in the pastoralist communities. John et al. (2022) stress this issue by noting that women in these communities encounter challenges in accessing maternal healthcare services because cultural beliefs prioritize home delivery and traditional methods over modern medical care. In support of this concept, Hussen et al. (2013) note that most pastoralists in Ethiopia prefer seeking medical opinion from traditional healers, which causes significant delay in seeking treatment, especially in the case of pulmonary tuberculosis. However, Okeibunor et al. (2013) found that the reliance on traditional healers for the healthcare needs of nomadic pastoralists in Nigeria occurs as they face significance limitations in accessing the required healthcare services.

Barasa and Virhia (2022) indicate that the interplay between dynamic and collective approaches to health, the broader cultural and social interpretation of health and diseases in people, the environment and animals play a significant role in decisions regarding seeking treatment. Ali and Woldearegai (2019) also noted that health-seeking behaviours, especially their decision on whether to visit the healthcare facility, have been motivated by the prevailing socio-cultural factors governing interpersonal relations in the community or in the family setup and how the treatment would impact other people within their communities and family. In promoting the importance of community members and the family, Atekem et al. (2022) indicated that adhering to healthy measures promoted through public health has been shaped by the overall attitude and acceptance of such measures by all in the community.

### 3.9 Improving Health Seeking Behaviours

The health-seeking behaviours in the context of the pastoralist communities can be enhanced by considering important aspects of health as remedies. Barasa and Virhia (2022), in this case, indicate that focusing on social and cultural factors such as gender, age, and notions of health is an important intervention in promoting positive health-seeking behaviour. Ahmed et al. (2018) also highlight the significance of proper dissemination of information about the availability of delivery services, which plays an important role in removing social and cultural barriers to increasing the use of delivery services for pastoralist women. Ali and Woldearegai (2019) indicate that positive health-seeking behaviours can be promoted through specific interventions, such as improved awareness of common diseases and the importance of diagnosis to reduce misconceptions. John et al. (2022) further indicate that outreach programs to modify cultural beliefs, enrolling more change agents in the community to promote maternal healthcare and integrating traditional and modern medical systems can be important in changing the myth around healthcare utilization. In the same case, Ahmed et al. (2018) stress the importance of education in order to enlighten community members on the importance of seeking medical opinions and services from qualified physicians. These authors noted that when women from the pastoralist communities are educated on the importance of delivery at the facility during their antenatal care (ANC) visits, they end up preferring facility delivery over home delivery (Ahmed et al., 2018). In support of these findings, Jackson and Hailemariam (2016) noted that women who are encouraged and informed on the importance of maternal health and other health issues during their ANC visits tend to be more proactive in seeking medical opinions about safe delivery. Tessema et al. (2019) also noted that when women are exposed to maternal education, they are more likely to understand the importance of medical treatment, especially with regard to vaccines for their children, portraying the importance of maternal education and public health awareness in changing negative health-seeking behaviours. Additionally, increasing access to quality services has been found to increase trust in healthcare facilities, which is an important factor in promoting positive health-seeking behaviour (Ali and Woldearegai, 2019).

Another important approach in improving health-seeking behaviours revolves around tailoring the intervention or healthcare services to the cultural and social norms of the communities. Kenny et al. (2021), in this case, indicate the significance of considering the preferences and fundamentals of the community, such as religion, norms, and power dynamics, when designing and implementing family planning interventions as interventions to improve the health of the population. Duale et al. (2023) found such improvement could be done through necessary training for skill development and capacity building among the healthcare staff. Byrne et al. (2016) indicate that capacity-building would be an important intervention because most pastoralist communities rely on and trust TBAs, who are, in most cases, not trained. For instance, Khogali et al. (2014) found that Self-administered treatments that are modified to resonate with the needs and the mobility nature of the pastoralist communities have been vital in improving their health while also changing their views regarding consulting medical officers for tuberculosis treatment.

Although cultural and social factors are seen as an important consideration in tailoring the health interventions to fit the preferences of pastoralists, Diaz (2017) indicates that these programmes need to consider the fact that in nature, pastoralists are always in movement and thus, mobile programmes would be effective in improving their health and promoting their health-seeking behaviours. Jackson and Hailemariam (2016), in this case, indicated that distance issues could be bridged by the provision of ambulances for transportation, especially in the case of emergency, because such an approach influences women’s decision to go to health centres for maternal health issues and delivery.

Another important outcome in promoting health-seeking behaviours among pastoralists has been linked to the use of community-based programmes. John et al. (2015), who assessed nomadic communities in Nigeria, noted that most of the interventions to improve treatment among these people were those done at the community level, steered by local leaders and other community volunteers. This was important in allowing the screening and treatment factors to be important aspects of the community. The same has been promoted by Okeibunor et al. (2013), who found that the community-directed intervention (CDI) strategy has great potential for improving access to health services among nomads, as it involves the community in the planning and implementation of different health interventions. Kaba and Mariam (2013) also highlight the importance of using people within the community because most of the pastoralist communities tend to ignore or fail to trust any information coming from outsiders.

## Chapter Four: Discussion

The results demonstrate a diverse and intricate situation concerning the health-seeking behaviours of nomadic pastoralists in low- and middle-income countries (LMICs). An essential determinant is the interaction between cultural attitudes and knowledge about health and sickness. Notably, the research conducted by Barasa and Virhia (2022), as well as Ag Ahmed et al. (2018), highlights the significance of cultural interpretations in understanding symptoms of sickness. Erroneous beliefs about illnesses such as fever, tuberculosis, and the normalisation of childbirth result in substantial delays in seeking medical assistance. These attitudes are firmly ingrained in conventional beliefs, whereby ailments are often ascribed to causes such as curses or witchcraft, hence further alienating the population from contemporary healthcare techniques. Traditional beliefs significantly hindered individuals from obtaining or pursuing medical aid. The primary concern was on the alignment between medical practices and traditional beliefs, which influenced the origins and manifestations of many illnesses (Ag-Ahmed et al., 2018). This aligns with the claims made by Cremers et al. (2013), which suggest that conventional notions regarding the origins of illnesses, such as witchcraft, could prompt the community to opt for traditional healing methods and refrain from seeking medical assistance or abandon treatment due to the perception that modern treatments are ineffective for such causes.

Moreover, gender roles and cultural conventions pose significant obstacles to the pastoralist communities’ ability to access and embrace contemporary care techniques and healthcare facilities. The research conducted by Kenny et al. (2021) and Henok and Takele (2017) demonstrates that there is a clear association between gender-based allocation of work and decision-making authority, which has a direct impact on individuals’ tendencies to seek healthcare. Men, who often hold the role of key decision-makers, may not place high importance on healthcare, particularly for women and children. This dynamic is further compounded by the inclination for traditional birth attendants over institutional treatment as a result of cultural beliefs and unease with male healthcare professionals. Moreover, a lack of knowledge regarding the causative agent and treatment, as well as traditional beliefs about the origins of diseases, can adversely affect patients’ attitudes towards seeking healthcare and taking preventive measures. Individuals who hold such beliefs are less likely to visit healthcare facilities (Sima et al., 2017). Consequently, this might lead to a significant delay in diagnosing and treating the condition, thus increasing the risk of disease transmission throughout the pastoralist community (Gebreegziabher et al., 2016). The absence of knowledge about the origin and management of illnesses has consistently been linked to subpar healthcare-seeking habits and delayed identification and treatment due to first consultations with traditional healers or non-medical practitioners (Engeda et al., 2016).

The presence of cultural variables and traditional beliefs not only hindered access to healthcare facilities but also contributed to the stigmatisation of seeking medical treatments, hence diminishing dependence on such services (Kenny et al., 2021). Pastoralists regarded tuberculosis (TB) as a highly significant and pressing issue within their community. They expressed fear and embarrassment at the prospect of being diagnosed with this disease, as the community tends to stigmatise TB patients (Sima et al., 2017). Individuals may experience personal stigma due to concerns about transmitting the disease, beliefs about the causes of the disease, or a desire to avoid discrimination within their community. On the other hand, community stigma may arise from the perceived risk of infection and the association between the disease and its perceived causes (Mushtaq et al., 2013).

The health-seeking behaviours of pastoralists are significantly influenced by their levels of education and expertise. The association between a lack of education and a restricted inclination to seek healthcare is apparent in the research conducted by Ali and Woldearegai (2019) as well as Hussen et al. (2013). Insufficient understanding of fundamental health information leads to misunderstandings and a widespread lack of confidence in contemporary healthcare systems. The findings of the current research indicate a notable lack of understanding among pastoralists on the causes, signs and symptoms, method of transmission, prevention, and treatment of tuberculosis (TB) (Sima et al., 2017). This signifies the limited dissemination of knowledge on the aetiology and management of illnesses within the pastoralist community. This may be attributed to the transitory nature of the population and the geographical features that provide challenges in reaching them via regular health education initiatives. This lack of comprehension worsens the unwillingness to seek timely and preventative medical treatment. Furthermore, the presence of institutional obstacles, such as the perception of disdain and mistreatment by healthcare practitioners, greatly discourages these groups from using the health services that are now accessible to them. This not only weakens the efficacy of health treatments but also adds to the continuous spread of avoidable illnesses, as shown in the instances of TB and malaria.

The hesitancy to participate in official healthcare systems has a significant effect on both the provision of health services and the overall health results of these populations. The research emphasises a detrimental cycle in which inadequate involvement with healthcare services as a result of cultural and information obstacles leads to worse health results, which then strengthens distrust and more disengagement. An example of this is the insufficient use of family planning and maternal healthcare services, as noted by Kenny et al. (2021). This not only impacts the health of individuals but also burdens the whole public health system. The difficulties experienced by healthcare practitioners in accessing these nomadic communities are worsened by the logistical and infrastructural hurdles, as pointed out by Duale et al. (2023) and Caulfield et al. (2016). This idea introduced the notion that distance might hinder individuals from accessing healthcare services or engaging with healthcare professionals. The primary factor influencing the decision to give birth at home was the considerable distance between the place of living and healthcare facilities (Ahmed et al., 2018). This element influenced the manner in which individuals obtained and their motivation to seek medical treatment from a healthcare institution. Therefore, improving the accessibility of healthcare institutions will enhance the likelihood of using the delivery services provided by these facilities. The pastoralists commonly sought assistance from traditional healers to address their ailments, perhaps due to limited access to healthcare within the pastoralist community (Sima et al., 2017; Hussen et al., 2013). The limited availability of healthcare services may be attributed to the absence of nearby health facilities, mostly owing to lengthy distances and inadequate transportation options. This issue is particularly prevalent among pastoralist communities since they are predominantly located in rural places (Esmael et al., 2013).

To tackle these issues, it is necessary to adopt a comprehensive strategy that recognises and incorporates the distinct cultural, social, and logistical elements of nomadic pastoralist groups. Promoting awareness and education on health and illness is of paramount importance. This entails implementing health education programmes that are culturally responsive and align with the community’s belief systems and behaviours. The use of indigenous languages and culturally significant symbols, together with the active participation of esteemed community members, may enhance comprehension and foster greater receptivity. Furthermore, healthcare treatments should be customised to correspond with the pastoralists’ lifestyles and social structures. One such approach is to recruit and educate community health professionals from these areas to act as intermediaries and reliable providers of healthcare knowledge and services.

Furthermore, enhancing healthcare accessibility is crucial for promoting health-seeking behaviours among marginalised groups. This applies not just to the physical accessibility but also to the price and cultural sensitivity of medical care. Mobile-based health clinics and telemedicine have the potential to serve as novel strategies for accessing these nomadic populations. Furthermore, community-directed interventions, as proposed by Okeibunor et al. (2013) and John et al. (2015), have the potential to effectively enhance community involvement and ownership of health programmes. To effectively enhance health-seeking behaviours among nomadic pastoralists in low- and middle-income countries (LMICs), it is necessary to tackle the intricate and interconnected obstacles of cultural attitudes, gaps in information, dynamics of gender, and institutional problems. An essential factor in improving health outcomes and the effectiveness of healthcare delivery in these distinct situations is adopting a cooperative, culturally aware, and community-oriented approach. Customising health treatments to suit the itinerant lifestyle and involving the community at every stage may result in substantial enhancements in health-seeking behaviours and overall health outcomes.

These results emphasise the need to develop customised tactics and health education interventions that are specially adapted for the pastoralist community. These interventions should aim to address the lack of information, negative attitudes, and healthcare-seeking behaviour associated with various medical disorders.

The research emphasised the need to formulate methods to effectively target nomadic pastoralists with preventive and therapeutic measures as nations progress towards eradicating prevalent illnesses. An analysis of the factors that affect decisions regarding delivery location can shed light on why numerous pastoralist women still choose to give birth at home. This knowledge can be utilised to shape policies and programmes that aim to enhance the percentage of deliveries taking place in healthcare facilities in this difficult context. By doing so, the health of mothers and newborns, as well as the outcomes of pastoralist communities, can be improved. The results provide a useful understanding of the intricate aspects that impact the decisions made by pastoralist women in regard to where they give birth. They emphasise the need to address socio-cultural attitudes and enhance the availability of considerate maternity care.

The study’s results closely correspond with the concepts outlined in Anderson’s Behavioural Model of Health Services Utilisation. This model, as described in the introduction chapter, suggests that the use of health services is impacted by three primary factors: predisposing features, enabling resources, and the desire for healthcare.

Predisposing characteristics refer to elements such as beliefs, attitudes, and knowledge that make persons more likely or less likely to seek healthcare services. The results emphasise the substantial influence of cultural ideas and misunderstandings on health, including the acceptance of certain symptoms as normal and the social disapproval of diseases such as tuberculosis. The hesitation to seek medical care is directly influenced by these attitudes, which are firmly established in traditional beliefs and practices. This phenomenon aligns with Anderson’s notion of predisposing traits.

Anderson defines enabling resources as the practical factors that determine one’s capacity to obtain healthcare, such as the presence, ease of access, and cost-effectiveness of healthcare services. The results indicate that nomadic pastoralists have significant obstacles in this field. Factors such as the division of labour based on gender, power dynamics in decision-making, and institutional impediments such as staff attitudes and challenges with hospital infrastructure restrict the accessibility and attractiveness of healthcare services. These issues align with the enabling resource aspect of Anderson’s paradigm, highlighting the need for resources that are not only readily available but also logistically and culturally accessible to these people. The term “Need for Healthcare” pertains to the perceived and assessed need for health services. The research results indicate that there is a lack of awareness of health issues among pastoralist groups, which may be attributed to their inadequate health knowledge and educational background. The incorrect understanding of health requirements results in delayed or absent efforts to seek healthcare, which corresponds to the ‘need’ aspect of Anderson’s model.

## Conclusion

The research sought to analyse and comprehend the health-seeking patterns of nomadic pastoralists, with a specific emphasis on the obstacles they face while trying to obtain high-quality healthcare facilities in LMICs. The thorough examination and evaluation of current literature provide a detailed and complex comprehension of these behaviours, providing insight into the many aspects that impact healthcare access and use among nomadic pastoralist populations. The results not only address but also provide a substantial contribution to the objectives of the research by thoroughly investigating the underlying difficulties.

The research highlights the intricate nature of health-seeking behaviours among nomadic pastoralists, which are shaped by a combination of cultural, educational, social, and institutional variables. The presence of negative beliefs and prejudices around diseases, which are strongly ingrained in the cultural framework of these communities, has emerged as a notable obstacle. These impressions, often resulting in the underestimation or misinterpretation of symptoms, are further exacerbated by conventional attitudes and practices that see some health problems as ordinary or not posing a significant concern. The cultural context significantly influences the attitude towards health and disease, often leading to a delay or avoidance of obtaining medical treatment.

Access to healthcare is further complicated by gender roles and cultural norms prevalent in these communities. The results emphasise the gendered aspects of healthcare access, including the authority of male community members in decision-making and the cultural incompatibility of women obtaining treatment from male healthcare practitioners. The aforementioned dynamics highlight the need to use gender-sensitive methods in the provision of healthcare and the formulation of policies.

The research emphasises the crucial significance of education and information in influencing health-seeking behaviours. Insufficient knowledge and comprehension about illnesses and their symptoms are factors that lead to the avoidance of healthcare. The lack of information in communities with low literacy levels highlights the need for educational initiatives that are culturally appropriate and easily available to nomadic pastoralists.

The challenges experienced by nomadic pastoralists in obtaining healthcare are worsened by institutional impediments, such as the attitudes of healthcare professionals and the insufficient healthcare infrastructure. The data suggest that these obstacles not only impede access but also impact the quality of treatment obtained. The prevailing scepticism about healthcare systems, driven by instances of disregard and perceived substandard treatment, underscores the urgent need to enhance healthcare provision and infrastructure in these areas.

The itinerant lifestyle inherently poses distinctive obstacles to obtaining healthcare. The nomadic nature of these tribes, along with the sometimes isolated areas they reside in, presents additional challenges in accessing healthcare services. Given the complex geography and logistical difficulties, it is crucial to come up with creative solutions, such as mobile health clinics, to guarantee the provision of healthcare in these always-changing environments. Financial limitations can have a pivotal impact on the ability to get healthcare services. The results emphasise the financial obstacles encountered by nomadic pastoralists, which often lead to the prioritisation of urgent economic needs above health concerns. The economic aspect of healthcare access is crucial, especially in low- and middle-income countries (LMICs) where limited resources are more noticeable.

### Strengths and Limitations

The incorporation of research from various areas throughout low- and middle-income countries (LMICs) enhances the analysis considerably. The wide geographical distribution of this study provides a comprehensive comprehension of the subject matter, taking into consideration the regional differences in cultural, social, and economic settings that impact individuals’ tendencies to seek healthcare. Additionally, Utilising a blend of qualitative, quantitative, and mixed-methods research methodologies improves the reliability of the results. Qualitative studies provide an in-depth understanding of the subjective experiences and cultural circumstances of pastoralist groups. Quantitative research provides quantifiable data that may be extrapolated to broader groups. Mixed-methods research combines both qualitative and quantitative approaches, providing a holistic perspective on the subject. The integration of several approaches and the inclusion of multiple geographical areas enables a more thorough comprehension of the complex nature of health-seeking behaviours among nomadic pastoralists in low- and middle-income countries (LMICs).

Due to a high concentration of research in Ethiopia and Kenya, there is a possibility of an overrepresentation of the unique concerns in these locations. This might possibly distort the general knowledge of health-seeking behaviours in all low- and middle-income countries (LMICs). Although the research primarily examines cultural, educational, and institutional obstacles, it adequately addresses other elements, such as political and policy-driven drivers. Furthermore, these components have a vital role in influencing individuals’ health-seeking behaviours.

### Recommendations

A key recommendation for boosting health-seeking habits is to conduct focused education and awareness efforts that specifically target misconceptions about illnesses and healthcare. These campaigns should use techniques and languages that are easily understood by the population. This may include prioritising health education activities at the community level, particularly in schools, to enhance health literacy starting from an early age. This research proposes the implementation of outreach activities to modify the cultural ideas of community members and enhance their comprehension of the significance of providing care to pregnant women.

One of the problems identified in the research is the presence of infrastructure impediments. Enhancing the healthcare infrastructure and improving its quality may play a crucial role in promoting use. Hence, enhancing the availability and accessibility of healthcare services in regions primarily populated by nomadic pastoralists is crucial. The findings of this research indicate that prioritising the enhancement of care standards at healthcare facilities and hospitals is essential. This entails allocating resources towards the establishment of mobile clinics and the use of telemedicine technologies, with the aim of delivering healthcare services to pastoralists. This may also include partnering with governments and NGOs to devise solutions that tackle the logistical and infrastructural obstacles in delivering healthcare to nomadic communities.

The research suggests the creation of healthcare programmes that are culturally sensitive and inclusive of the beliefs and practices of nomadic pastoralist groups. Moreover, the government should contemplate the incorporation of ancient medical systems into contemporary medical systems. This would furthermore include actively involving local leaders and community people in the planning and execution of health interventions to ensure that they are appropriate to the specific circumstances and more likely to be embraced.

An additional crucial factor to consider is the instruction of indigenous members of the community as healthcare practitioners, which serves to overcome cultural and linguistic barriers and fosters trust among pastoralists. This would also include promoting the enlistment of more female healthcare practitioners to address the specific requirements of women in these areas while acknowledging and valuing gender dynamics.

Public health personnel must also prioritise policy advocacy and reform as a crucial factor in boosting health-seeking behaviours. Specifically, supporting policies that prioritise the distinct healthcare requirements of nomadic pastoralists, such as providing enough financing and allocating resources, is a crucial factor in encouraging favourable health practices.

### Future Research

Future studies should concentrate on the economic obstacles to health access, such as healthcare services and transportation costs, and how these influence health-seeking behaviours. Future studies should look at how technology, such as mobile health applications and telemedicine, might improve healthcare access and monitoring for nomadic people. This covers the potential of digital technologies for data collecting and health monitoring in nomadic groups.

## Data Availability

All data produced in the present work are contained in the manuscript

## References

Abuduxike, G., Aşut, Ö., Vaizoğlu, S.A. and Cali, S., 2020. Health-seeking behaviours and its determinants: a facility-based cross-sectional study in the Turkish Republic of Northern Cyprus. International journal of health policy and management, 9(6), p.240.

Ahmed, M., Demissie, M., Medhanyie, A.A., Worku, A. and Berhane, Y., 2018. utilization of institutional delivery Services in a Predominantly Pastoralist Community of Northeast Ethiopia. Ethiopian journal of health sciences, 28(4).

Ag Ahmed, M.A., Hamelin-Brabant, L. and Gagnon, M.P., 2018. Sociocultural determinants of nomadic women’s utilization of assisted childbirth in Gossi, Mali: a qualitative study. BMC Pregnancy and childbirth, 18, pp.1–14.

Ali, M., J.P. Cordero, F. Khan, and R. Folz. 2019. ‘Leaving no one behind’: A scoping review on the provision of sexual and reproductive health care to nomadic populations. BMC Women’s Health 19: 161.

Ali, S.S. and Woldearegai, B.T., 2019. Health-seeking behaviour of afar pastoral community. Int J Eng Adv Technol, 8(5), pp.292–6.

Amegbor, P.M., 2014. Health seeking behaviour in Asikuma-Odoben-Brakwa district: A pluralistic health perspective.

Andersen, R.M., 1995. Revisiting the behavioural model and access to medical care: does it matter? Journal of Health and Social Behavior, pp.1–10.

Andersen, R.M., Rice, T.H. and Kominski, G.F., 2011. Changing the US health care system: Key issues in health services policy and management. John Wiley & Sons.

Anthonj, C., Giovannini, P. and Kistemann, T., 2019. Coping with ill-health: health care facility, chemist or medicinal plants? Health-seeking behaviour in a Kenyan wetland. BMC International Health and Human Rights, 19, pp.1–14.

Atekem, K., Dixon, R., Nditanchou, R., Makia, C.M., Ntsinda, M., Basnet, S. and Schmidt, E., 2022. Reach and utility of COVID-19 information and preventive measures for nomadic populations in Massangam, West region of Cameroon. The American Journal of Tropical Medicine and Hygiene, 106(5), p.1491.

Ayalew, H.G., Liyew, A.M., Tessema, Z.T., Worku, M.G., Tesema, G.A., Alamneh, T.S., Teshale, A.B., Yeshaw, Y. and Alem, A.Z., 2022. Spatial variation and factors associated with home delivery after ANC visit in Ethiopia; spatial and multilevel analysis. Plos one, 17(8), p.e0272849.

Barasa, V. and Virhia, J., 2022. Using Intersectionality to Identify gendered Barriers to Health-seeking for Febrile Illness in Agro-pastoralist settings in Tanzania. Frontiers in Global Women’s Health, 2, p.746402.

Byrne, A., Caulfield, T., Onyo, P., Nyagero, J., Morgan, A., Nduba, J. and Kermode, M., 2016. Community and provider perceptions of traditional and skilled birth attendants providing maternal health care for pastoralist communities in Kenya: a qualitative study. BMC pregnancy and childbirth, 16, pp.1–12.

Cattaneo, V., 2019. Key influential factors impacting access to primary healthcare among nomadic communities.

Caulfield, T., Onyo, P., Byrne, A., Nduba, J., Nyagero, J., Morgan, A. and Kermode, M., 2016. Factors influencing place of delivery for pastoralist women in Kenya: a qualitative study. BMC Women’s Health, 16, pp.1–11.

Cremers, A.L., Janssen, S., Huson, M.A., Bikene, G., Bélard, S., Gerrets, R.P. and Grobusch, M.P., 2013. Perceptions, health care seeking behaviour and implementation of a tuberculosis control programme in Lambaréné, Gabon. Public Health Action, 3(4), pp.328–332.

Da Silva, R.B., Contandriopoulos, A.P., Pineault, R. and Tousignant, P., 2011. A global approach to evaluation of health services utilization: concepts and measures. Healthcare Policy, 6(4), p.e106.

Diaz, M., 2017. Factors associated with non-vaccination among nomadic pastoralists and under-vaccination among settled pastoralists in Lagdera, Kenya.

Duale, H.A., Farah, A., Salad, A., Gele, S. and Gele, A., 2023. Constraints to maternal healthcare access among pastoral communities in the Darussalam area of Mudug region, Somalia “a qualitative study”. Frontiers in Public Health, 11.

El Shiekh, B. and van der Kwaak, A., 2015. Factors influencing the utilization of maternal health care services by nomads in Sudan. Pastoralism, 5(1), p.23.

Engeda, E.H., Dachew, B.A., Kassa Woreta, H., Mekonnen Kelkay, M. and Ashenafie, T.D., 2016. Health seeking behaviour and associated factors among pulmonary tuberculosis suspects in lay Armachiho District, Northwest Ethiopia: A Community-Based Study. Tuberculosis research and treatment, 2016.

Esmael, A., Ali, I., Agonafir, M., Desale, A., Yaregal, Z. and Desta, K., 2013. Assessment of patients’ knowledge, attitude, and practice regarding pulmonary tuberculosis in eastern Amhara regional state, Ethiopia: a cross-sectional study. The American Journal of Tropical Medicine and Hygiene, 88(4), p.785.

Gammino, V.M., Diaz, M.R., Pallas, S.W., Greenleaf, A.R. and Kurnit, M.R., 2020. Health services uptake among nomadic pastoralist populations in Africa: a systematic review of the literature. PLoS Neglected Tropical Diseases, 14(7), p.e0008474.

Gebreegziabher, S.B., Bjune, G.A. and Yimer, S.A., 2016. Patients and health system’s delays in the diagnosis and treatment of new pulmonary tuberculosis patients in West Gojjam Zone, Northwest Ethiopia: a cross-sectional study. BMC infectious diseases, 16, pp.1–13.

Hanelt, A., Bohnsack, R., Marz, D. and Antunes Marante, C., 2021. A systematic review of the literature on digital transformation: Insights and implications for strategy and organizational change. Journal of Management Studies, 58(5), pp.1159–1197.

Henok, A. and Takele, E., 2017. Assessment of barriers to reproductive health service utilization among Bench Maji Zone Pastoralist Communities. Ethiopian Journal of Health Sciences, 27(5), pp.523–530.

Holde, G.E., Baker, S.R. and Jönsson, B., 2018. Periodontitis and quality of life: What is the role of socioeconomic status, sense of coherence, dental service use and oral health practices? An exploratory theory-guided analysis of a Norwegian population. Journal of Clinical Periodontology, 45(7), pp.768–779.

Hussen, A., Biadgilign, S., Tessema, F., Mohammed, S., Deribe, K. and Deribew, A., 2013. Treatment delay among pulmonary tuberculosis patients in pastoralist communities in Bale Zone, Southeast Ethiopia. BMC research notes, 5, pp.1–10.

Jackson, R. and Hailemariam, A., 2016. The role of health extension workers in linking pregnant women with health facilities for delivery in rural and pastoralist areas of Ethiopia. Ethiopian Journal of Health Sciences, 26(5), pp.471–478.

Jillo, J.A., Ofware, P.O., Njuguna, S. and Mwaura-Tenambergen, W., 2015. Effectiveness of Ng’adakarin Bamocha model in improving access to ante-natal and delivery services among nomadic pastoralist communities of Turkana West and Turkana North Sub-Counties of Kenya. The Pan African Medical Journal, 20.

John, S., Gidado, M., Dahiru, T., Fanning, A., Codlin, A.J. and Creswell, J., 2015. Tuberculosis among nomads in Adamawa, Nigeria: outcomes from two years of active case finding. The International Journal of Tuberculosis and Lung Disease, 19(4), pp.463–468.

John, M., Vundi, N. and Gichuhi, D., 2022. Cultural beliefs influencing access to maternal healthcare services in East Pokot Pastoral communities, Baringo County, Kenya. International Journal of Research in Business and Social Science (2147-4478), 11(7), pp.331–339.

Kaba, M. and Mariam, D.H., 2013. The state of HIV awareness after three decades of intervention in Ethiopia: the case of the Borana pastoral community in Southern Ethiopia. Ethiopian Journal of Health Development, 26(1), pp.9–15.

Kenny, L., Lokot, M., Bhatia, A., Hassan, R., Pyror, S., Dagadu, N.A., Aden, A., Shariff, A., Bacchus, L.J., Hossain, M. and Cislaghi, B., 2022. Gender norms and family planning amongst pastoralists in Kenya: a qualitative study in Wajir and Mandera. Sexual and reproductive health matters, 30(1), p.2135736.

Kenya National Bureau of Statistics. 2015. Kenya Demographic and Health Survey 2014. Nairobi: Kenya National Bureau of Statistics.

Khogali, M., Zachariah, R., Reid, T., Alipon, S.C., Zimble, S., Mahama, G., Etienne, W., Veerman, R., Dahmane, A., Weyeyso, T. and Hassan, A., 2014. Self-administered treatment for tuberculosis among pastoralists in rural Ethiopia: how well does it work? International Health, 6(2), pp.112–117.

Lau, R., Stevenson, F., Ong, B.N., Dziedzic, K., Treweek, S., Eldridge, S., Everitt, H., Kennedy, A., Qureshi, N., Rogers, A. and Peacock, R., 2015. Achieving change in primary care—causes of the evidence to practice gap: systematic reviews of reviews. Implementation Science, 11(1), pp.1–39.

Lechthaler, F., M.F. Abakar, E. Schelling, J. Hattendorf, B. Ouedraogo, D.D. Moto, and J. Zinsstag. 2018. Bottlenecks in the provision of antenatal care: Rural settled and mobile pastoralist communities in Chad. Tropical Medicine & International Health 23: 1033– 1044.

Lô, A., Tall-Dia, A., Bonfoh, B. and Schelling, E., 2016. Tuberculosis among transhumant pastoralist and settled communities of south-eastern Mauritania. Global health action, 9(1), p.30334.

Maro, G, Nguura, P, Umer, J, Gitimu, A, Haile, F, Kawai, D, and Lukmay, K. 2012. Understanding nomadic realities-Case studies on sexual and reproductive health and rights in Eastern Africa. In, eds. A. Van Der Kwaak et al. Amsterdam: AMREF/KIT.

Montavon, A., Jean-Richard, V., Bechir, M., Daugla, D.M., Abdoulaye, M., Bongo Naré, R.N., Diguimbaye-Djaibé, C., Alfarouk, I.O., Schelling, E., Wyss, K. and Tanner, M., 2013. Health of mobile pastoralists in the Sahel–assessment of 15 years of research and development. Tropical Medicine & International Health, 18(9), pp.1044–1052.

Munn, Z., Peters, M.D., Stern, C., Tufanaru, C., McArthur, A. and Aromataris, E., 2018. Systematic review or scoping review? Guidance for authors when choosing between a systematic or scoping review approach. BMC Medical Research Methodology, 18, pp.1–7.

Muriithi, F.G., Banke-Thomas, A., Gakuo, R., Pope, K., Coomarasamy, A. and Gallos, I.D., 2022. Individual, health facility and wider health system factors contributing to maternal deaths in Africa: A scoping review. PLOS global public health, 2(7), p.e0000385.

Mushtaq, M.U., Shahid, U., Abdullah, H.M., Saeed, A., Omer, F., Shad, M.A., Siddiqui, A.M. and Akram, J., 2013. Urban-rural inequities in knowledge, attitudes and practices regarding tuberculosis in two districts of Pakistan’s Punjab province. International Journal for Equity in Health, 10, pp.1–9.

Ngwakongnwi, E., 2017. Measuring health services utilization in ethnic populations: Ethnicity and choice of frameworks. Public Health Open J, 2(2), pp.53–58.

Okeibunor, J.C., Onyeneho, N.G., Nwaorgu, O.C., I’Aronu, N., Okoye, I., Iremeka, F.U. and Sommerfeld, J., 2013. Prospects of using community-directed intervention strategy in delivering health services among Fulani Nomads in Enugu State, Nigeria. International journal for equity in health, 12(1), pp.1–17.

Pertet, A.M., Kaseje, D., Otieno-Odawa, C.F., Kirika, L., Wanjala, C., Ochieng, J., Jaoko, M., Otieno, W. and Odindo, D., 2018. Under vaccination of children among Maasai nomadic pastoralists in Kenya: is the issue geographic mobility, social demographics or missed opportunities? BMC Public Health, 18(1), pp.1–9.

Peters, M.D., Marnie, C., Tricco, A.C., Pollock, D., Munn, Z., Alexander, L., McInerney, P., Godfrey, C.M. and Khalil, H., 2021. Updated methodological guidance for the conduct of scoping reviews. JBI evidence implementation, 19(1), pp.3–10.

Peterson, J., Pearce, P.F., Ferguson, L.A. and Langford, C.A., 2017. Understanding scoping reviews: Definition, purpose, and process. Journal of the American Association of Nurse Practitioners, 29(1), pp.12–16.

Pham, M.T., Rajić, A., Greig, J.D., Sargeant, J.M., Papadopoulos, A. and McEwen, S.A., 2014. A scoping review of scoping reviews: advancing the approach and enhancing the consistency. Research synthesis methods, 5(4), pp.371–385.

Ringo, J.J., Bengesi, K.M. and Mbago, M.C., 2018. Access and challenges of health facilities amongst agro-pastoralist communities in Handeni District, Tanzania. Journal of Population and Social Studies [JPSS], 26(1), pp.53–67.

Seck, M.C., Thwing, J., Fall, F.B., Gomis, J.F., Deme, A., Ndiaye, Y.D., Daniels, R., Volkman, S.K., Ndiop, M., Ba, M. and Ndiaye, D., 2017. Malaria prevalence, prevention and treatment seeking practices among nomadic pastoralists in northern Senegal. Malaria journal, 16, pp.1–11.

Shayo, E.H., Rumisha, S.F., Mlozi, M.R., Bwana, V.M., Mayala, B.K., Malima, R.C., Mlacha, T. and Mboera, L.E., 2015. Social determinants of malaria and health care seeking patterns among rice farming and pastoral communities in Kilosa District in central Tanzania. Acta tropica, 144, pp.41–49.

Sima, B.T., Belachew, T. and Abebe, F., 2017. Knowledge, attitude and perceived stigma towards tuberculosis among pastoralists: Do they differ from sedentary communities? A comparative cross-sectional study. PloS one, 12(7), p.e0181032.

Tessema, F., Bisrat, F., Kidane, L., Assres, M., Tadesse, T. and Asegedew, B., 2019. Improvements in polio vaccination status and knowledge about polio vaccination in the CORE Group Polio Project implementation areas in pastoralist and semi-pastoralist regions in Ethiopia. The American Journal of Tropical Medicine and Hygiene, 101(4 Suppl), p.52.

Travers, J.L., Hirschman, K.B. and Naylor, M.D., 2020. Adapting Andersen’s expanded behavioural model of health services use to include older adults receiving long-term services and supports. BMC geriatrics, 20(1), pp.1–16.

Wako, W.G., and D.H. Kassa. 2017. Institutional delivery service utilization and associated factors among women of reproductive age in the mobile pastoral community of the Liban District in Guji Zone, Oromia, Southern Ethiopia: A cross-sectional study. BMC Pregnancy and Childbirth 17: 1–10.

WHO. 2018. A Vision for Primary Health Care in the 21st Century: Towards Universal Health Coverage and the Sustainable Development Goals. WHO and UNICEF New York. https://www.who.int/docs/default-source/primary-health/vision.pdf.

WHO. 2019. World Health Day 2019: Campaign Essentials. https://www.who.int/campaigns/world-health-day/world-health-day-2019.

Wild, H., Glowacki, L., Maples, S., Mejía-Guevara, I., Krystosik, A., Bonds, M.H., Hiruy, A., LaBeaud, A.D. and Barry, M., 2019. Making pastoralists count: geospatial methods for the health surveillance of nomadic populations. The American Journal of Tropical Medicine and Hygiene, 101(3), p.661.

Wulifan, J.K., S. Brenner, A. Jahn, and M. De Allegri. 2016. A scoping review on determinants of unmet need for family planning among women of reproductive age in low and middle-income countries. BMC Women’s Health 16: 2.

Wulifan, J.K., Dordah, A.D. and Sumankuuro, J., 2022. Nomadic pastoralists’ experience accessing reproductive and maternal healthcare services in low and middle-income countries: A contextual scoping review. Pastoralism, 12(1), p.47.

